# Safety and efficacy of left bundle branch area pacing compared with right ventricular pacing in patients with bradyarrhythmia and conduction system disorders: a systematic review and meta-analysis

**DOI:** 10.1101/2023.02.09.23285675

**Authors:** Georgios Leventopoulos, Christoforos K. Travlos, Konstantinos N. Aronis, Virginia Anagnostopoulou, Panagiotis Patrinos, Aggeliki Papageorgiou, Angelos Perperis, Chris P. Gale, Periklis Davlouros

## Abstract

**Background:** Right Ventricular Pacing (RVP) may have detrimental effects in ventricular function. Left Bundle Branch Area Pacing (LBBAP) is a new pacing strategy that appears to have better results. The aim of this systematic review and meta-analysis is to compare the safety and efficacy of LBBAP vs RVP in patients with bradyarrhythmia and conduction system disorders.

**Methods:** Medline, Embase and Pubmed databases were searched for studies comparing LBBAP with RVP. Outcomes were all-cause mortality, atrial fibrillation (AF) occurrence, heart failure hospitalizations (HFH) and complications. QRS duration, mechanical synchrony and LVEF changes were also assessed. Pairwise meta-analysis was conducted using random and fixed effects models.

**Results:** Twenty-five trials with 4250 patients (2127 LBBAP) were included in the analysis. LBBAP was associated with lower risk for HFH (RR:0.33, CI 95%:0.21 to 0.50; *p*<0.001), all-cause mortality (RR:0.52 CI 95%:0.34 to 0.80; *p*=0.003), and AF occurrence (RR:0.43 CI 95%:0.27 to 0.68; *p*<0.001) than RVP. Lead related complications were not different between the two groups (*p=*0.780). QRSd was shorter in the LBBAP group at follow-up (WMD: -32.20 msec, CI 95%: -40.70 to -23.71; *p*<0.001) and LBBAP achieved better intraventricular mechanical synchrony than RVP (SMD: -1.77, CI 95%: -2.45 to -1.09; *p*<0.001). LBBAP had similar pacing thresholds (*p*=0.860) and higher R wave amplitudes (*p*=0.009) than RVP.

**Conclusions:** LBBAP has better clinical outcomes, preserves ventricular electrical and mechanical synchrony and has excellent pacing parameters, with no difference in complications compared to RVP.

**Clinical Perspective:** *What is known:*

- Left bundle branch area pacing (LBBAP) is a method of conduction system pacing with higher procedural success rate and less limitations compared to His bundle pacing (HBP).
- Right ventricular pacing (RVP) causes electromechanical dyssynchrony, which may result in left ventricular systolic dysfunction in some patients.

*What the study adds:*

- We examined in a systematic review and meta-analysis whether there was a difference in clinical outcomes, electromechanical synchronization, and safety between LBBAP and RVP in patients with bradyarrhythmia and conduction system disorders.

## 1. Introduction

Right ventricular pacing (RVP) comprised of right ventricular apical pacing (RVAP), right ventricular septal pacing (RVSP) and right ventricular outflow tract pacing (RVOP) is well established as the widely accepted pacing method.^1,2^ RVP presents the advantages of easy implantation, good pacing parameters and low rate of lead dislodgement. However, its effects on the ventricles can be detrimental causing electrical and mechanical dyssynchrony which can lead to heart failure hospitalization (HFH) with rates up to 9.6%, atrial fibrillation (AF) in 21-24% of patients, and pacing induced cardiomyopathy in 19.5% of patients.^3-7^

To address the need for more physiological pacing without the negative effects of RVP in cardiac function, Deshmukh et al. was the first to introduce His-bundle pacing (HBP) in humans, in 2000.^8^ This appeared to be a promising new technique ensuring rapid and synchronized contraction of the left and right ventricle and providing electrical synchrony by directly engaging the His–Purkinje system of the heart.^9^ Nevertheless, HBP has limitations that have prevented it from becoming a wide-spread alternative to RVP. Implantation may be challenging, and the high pacing threshold, the increased percentage of lead revision, and the low success rates, specifically in patients with infranodal block and bundle branch block (BBB) make its use limited.^10,11^

Left bundle branch area pacing (LBBAP) has emerged as an alternative to achieve conduction system pacing.^12^ Huang et al., first demonstrated the direct capture of left bundle (LB) and achieved synchronized activation of ventricles by placing the ventricular lead deep inside the intraventricular septum.^13^ Thus, LBBAP emerged as a viable alternative to RVP with the advantage of overcoming the clinical difficulties of HBP. Several studies have evaluated the effects of LBBAP in cardiac function. However, a large study comparing the effects of LBBAP to RVP, specifically in patients with bradyarrhythmia and conduction system disorders is still lacking. To our knowledge, this is the first systematic review and meta-analysis that aims to compare the safety and efficacy of LBBAP with RVP in patients with bradycardia and conduction system disorders.

## 2. Methods

This systematic review and meta-analysis was performed in accordance with the Preferred Reporting Items for Systematic Reviews and Meta-Analyses (PRISMA) guidelines (supplementary material).^14^ This study was registered at the Prospective International Register of Systematic Reviews (PROSPERO, registration number CRD42022315046). All data used and analyses performed in this systematic review and meta-analysis were based in previously published studies.

### 2.1 Search strategy

The research question was structured using the PICOT (Population, Intervention, Comparison, Outcomes, Type of studies) question frame (supplementary material). We searched Medline, Embase (via Ovid framework) and PubMed databases for studies comparing LBBAP with RVP from inception through November 10, 2022. The full search strategy is provided in supplementary material. References from included studies were manually searched for potentially relevant publications not identified from initial search. Endnote’s duplicate detection function was used to detect duplicates.

### 2.2 Study selection

Clinical studies were eligible if they met the following inclusion criteria: (1) randomized controlled trials (RCTs) and observational studies that compared an LBBAP group with an RVP group in bradycardia or conduction system disorders; (2) studies comparing clinical outcomes, complications, pacing parameters, echocardiographic changes, electrophysiology characteristics, between LBBAP and RVP in patients with a left ventricular ejection fraction (LVEF) >35%; (3) RVP group included either right ventricular apical pacing, right ventricular septal pacing, or right ventricular outflow tract pacing; (4) articles published in peer-reviewed journals with full text available.

We excluded: Animal studies, case reports, review articles, editorials, letters, congress abstracts, studies in individuals aged <18 years, studies including <10 participants.

Records were uploaded to a systematic review web application (Rayyan, Qatar Computing Research Institute).^15^ Two independent investigators (CT and VA) screened articles for inclusion. Disagreements were resolved by discussion and if consensus couldn’t be reached a third senior investigator (GL) was consulted.

### 2.3 Outcomes

Outcomes included:

a. Clinical outcomes such as HFH, AF occurrence and all-cause mortality
b. Lead related complications (lead dislodgement, lead perforation during the procedure, late lead perforation)
c. Ventricular electrical synchrony, assessed by paced QRS duration (QRSd), and stim-LVAT (left ventricle activation time)
d. Left ventricular (LV) mechanical synchrony which was assessed by comparing intraventricular synchrony and interventricular synchrony separately. We included studies that measured the dyssynchrony between different segments of the left ventricle using SPWMD (septal to posterior wall motion delay), TS-12-SD, phase analysis using phase bandwith, Tmsv16-SD/R-R and studies that evaluated the presence of dyssynchrony between the two ventricles, using the interventricular mechanical delay (IVMD).
e. Left ventricular systolic function assessed by LVEF and left ventricle end-diastolic diameter (LVEDD)
f. Pacing parameters including pacing threshold, ventricular impedance, R wave amplitude
g. Procedural characteristics including procedural duration, fluoroscopy time, procedural success rate, probability of recording left bundle brunch (LBB) potential and correction of BBB.

For each outcome data were collected at baseline, immediately after the procedure and at the longest follow-up if available. We performed analyses to compare LBBAP with pre-LBBAP implantation characteristics, as well as LBBAP versus RVP. For each study we also collected data concerning study type, number of patients and indication for pacing in each group, patient baseline characteristics, type of lead used and follow-up duration.

### 2.4 Data collection and extraction

Two independent investigators (CT and VA) reviewed the full text and supplement of included studies and extracted data in an excel spreadsheet using the same protocol. Discrepancies were resolved by discussion and by consulting a third investigator (GL). If data were not available in the text, then we translated graphic information to numerical data by using a suitable imaging software.^16^ The use of this software results in a small absolute difference from the true values and excellent consistency. According to Burda et al. ^17^ it can be used in systematic reviews as it has a small margin of error. Data concerning study type, number of patients, baseline characteristics and rate of success are presented in **Table 1**. The sample mean and standard deviation was extracted for each data type. In case that results were reported as median and interquartile range we converted them in sample mean and standard deviation by using the Wan’s et al. method.^18^

**Table 1:**
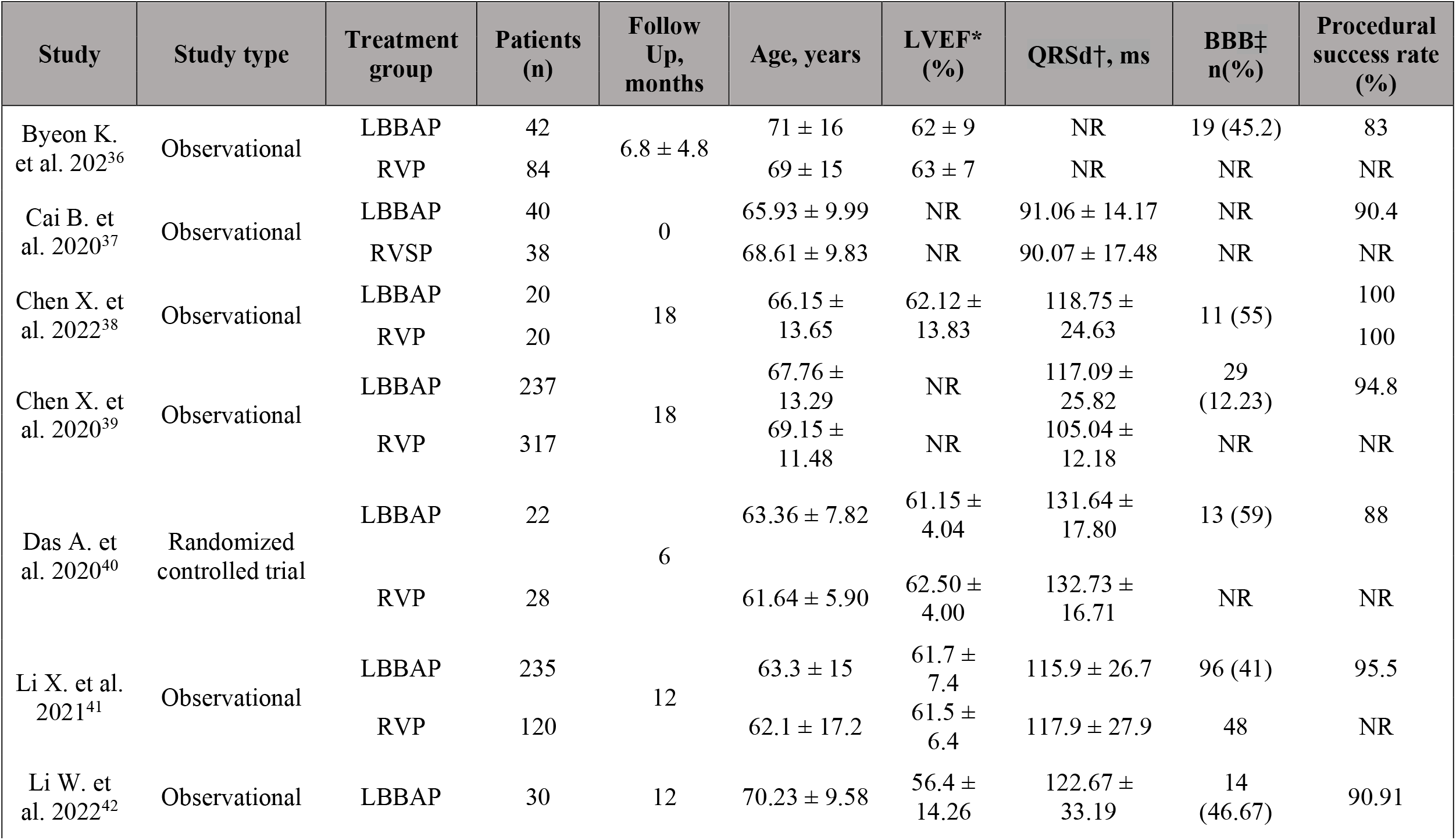
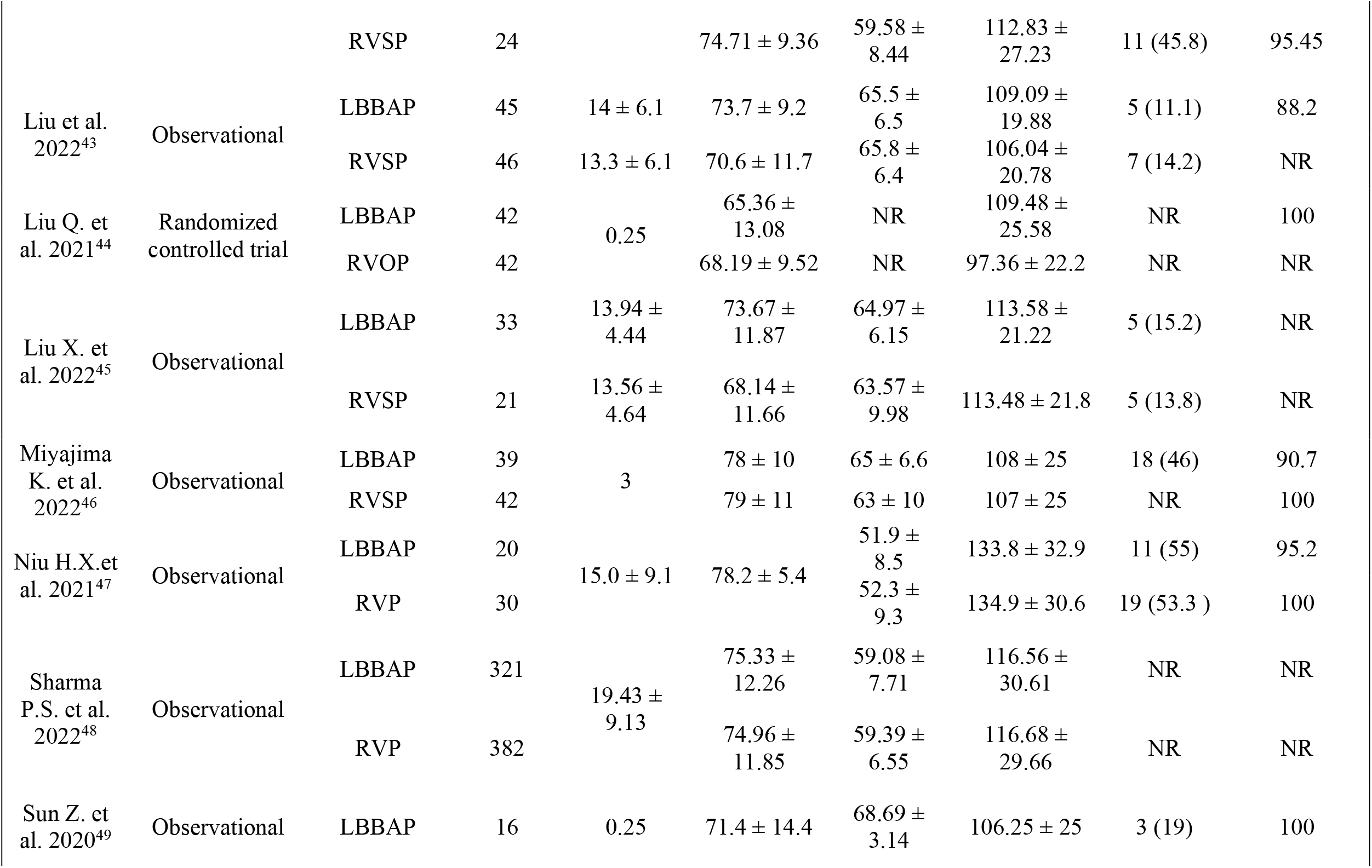
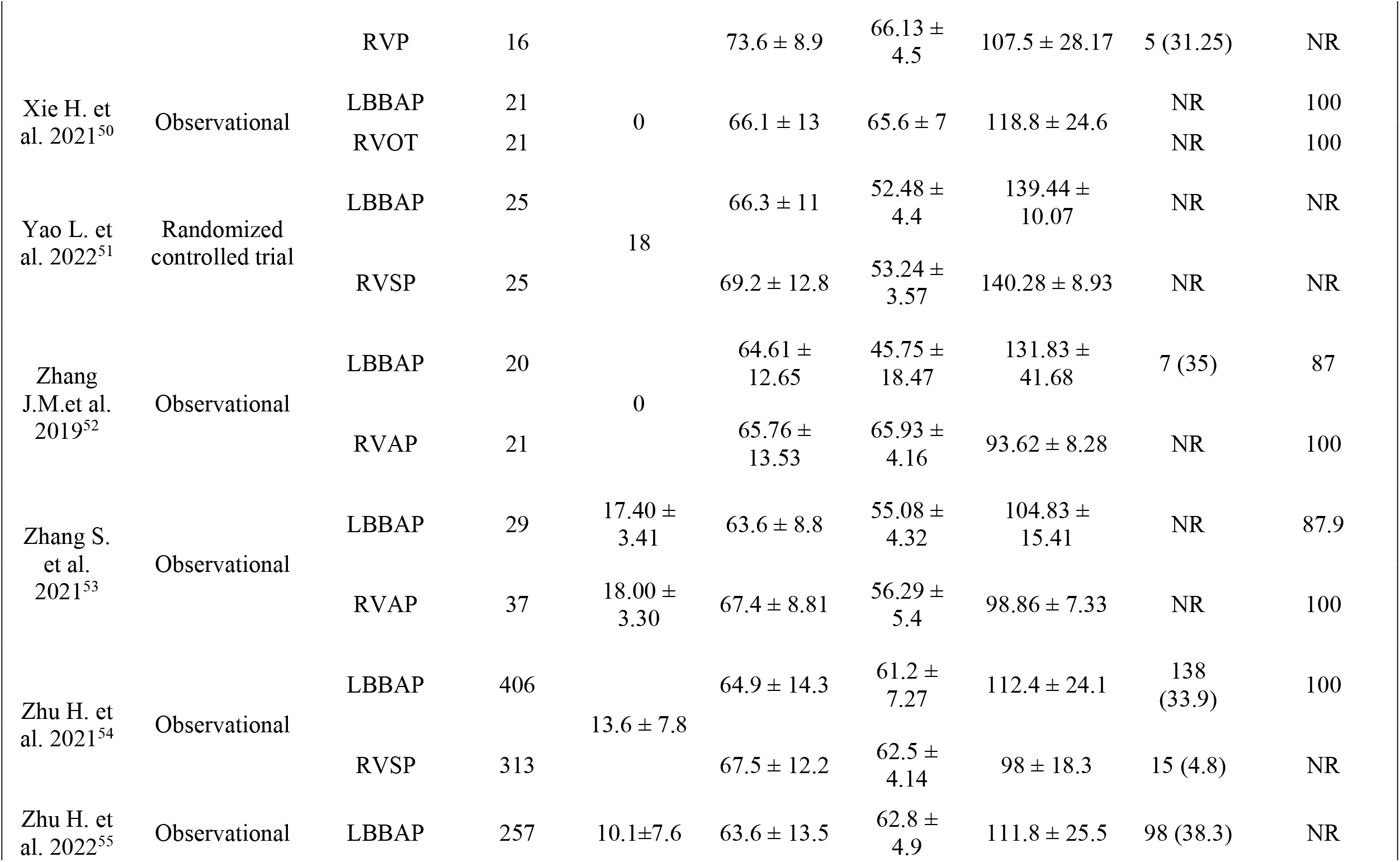
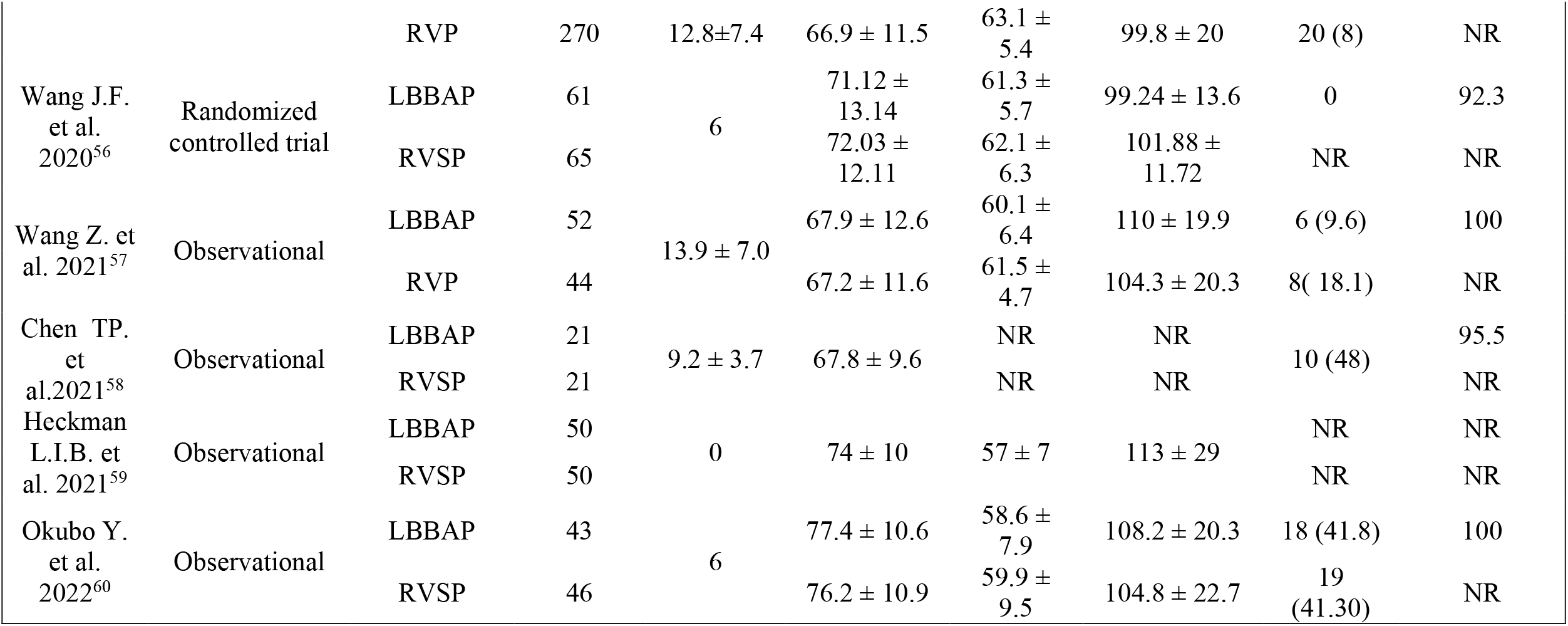
Baseline and procedural characteristics of included studies. * LVEF: left ventricular ejection fraction, † QRSd: QRS duration, ‡ BBB: bundle branch block

### 2.5 Quality assessment

We assessed the quality of included studies by using the Cochrane Risk of Bias 2 assessment tool (ROB 2.0) for RCTs and the Newcastle–Ottawa scale (NOS) for observational studies.^19,20^ The NOS uses a star system (0–9) to evaluate studies. If a study had a NOS ≥7 we considered it as a study of good quality.

### 2.6 Statistical analysis

The data for each outcome of interest from the included studies, were pooled (mean value, standard deviation and sample size for continuous variables and number of events and sample size for dichotomous variables), to compare the outcomes between LBBAP with pre-LBBAP implantation characteristics and LBBAP vs RVP. The effect measure for continuous variables was weighed mean difference (WMD) if the outcome was measured the same way in all studies and standard mean difference (SMD) if the method of measurement varied between studies. Dichotomous variables were reported as risk ratios (RR) and 95% confidence intervals (CIs) were used both for continuous and dichotomous outcomes. The between-study heterogeneity was assessed using Cochran’s Q test and Higgins I^2^ statistic. 0%, <25%, 25% to 49%, and >50% denoted no, low, moderate, and high heterogeneity, respectively. If I^2^ value was less than 50%, a fixed-effects (Mantel–Haenszel) model was adopted. Otherwise, a random-effects (DerSimonian-Laird) model was used considering the substantial heterogeneity. In cases of statistical heterogeneity, subgroup analysis or sensitivity analyses were used. Sensitivity analyses were performed by removing one study at one time to explore the consistency of the results (“leave-one-out sensitivity analysis”). The risk of potential publication bias was assessed with Funnel plots and Egger’s test ^21^ in outcome comparisons that included more than 8 studies. All p values were two-sided, with p<0.05 considered as significant. All statistical analyses were performed using RevMan 5.4 ^22^ and Jamovi software ^23^.

## 3. Results

### 3.1 Study and data selection

In total 1318 studies were retrieved (94 from Medline, 128 from Embase and 1096 from PubMed). After duplicates removal 1138 studies remained for screening and 53 studies were included for full text review. Finally, 25 studies were included in the systematic review and meta-analysis. Full study selection process is shown in **Figure 1**. No additional relevant studies were found from manual search of references lists.

**Figure 1:**
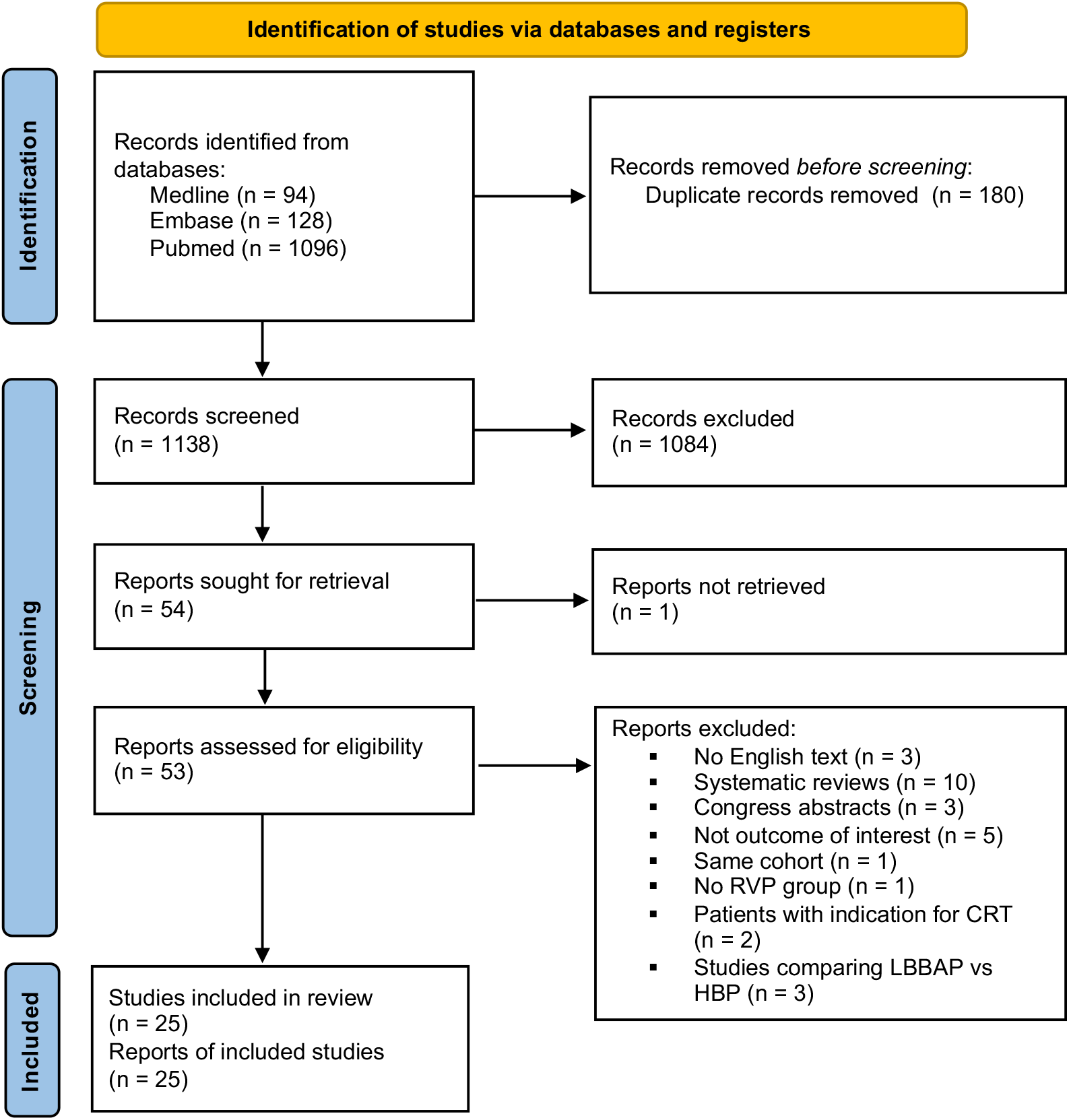
Flow diagram of literature search.

### 3.2 Characteristics of included studies

We included 21 observational studies and four RCTs; all compared LBBAP with RVP. The LBBAP group comprised patients undergoing LBBP, LBBAP and LVSP depending on which pacing method was used in each individual study. RVP group was comprised of RVAP, RVSP and RVOP depending on which pacing method was used in each individual study. In the RVP group, ten studies included only RVSP, eight studies included RVSP or RVAP without individual data for each subgroup (only data for the whole RVP group were available), two studies included RVSP and RVAP group with individual data available for each subgroup, two studies included only RVAP, one study RVOP, one study RVAP and RVOP with individual data available for each subgroup and one study included RVP without clarifying the specific pacing position. In the studies where RVP group population was separated into RVSP and RVAP or RVOP and RVAP subgroups, we collected and analyzed data only for the RVSP and RVOP subgroup, respectively. Considering that RVSP and RVOP are closer to physiological pacing than RVAP, less bias would be introduced that way.

Patient baseline and procedural characteristics are presented in **Table 1**. A total of 4250 individuals were enrolled in these 25 trials (2127 in the LBBAP group and 2123 in the RVP group). In all studies, the pacing indications were either sinus node dysfunction (SND), atrioventricular block (AVB) or atrial fibrillation (AF) with a slow ventricular rate. The mean follow-up (FU) duration was 11.2 ± 6.1 months with a range from 0 to 29 months with no difference between LBBAP and RVP group. The mean age of the participants was 70.4 ± 11.6 years and the mean LVEF was 60.58 ± 7.24 %. The mean success rate of LBBAP in the included studies was 93.6%, and the average probability of recording LBB potential was 61.8%. In 24 studies, the SelectSecure system (model 3830 lead, 69 cm; C315 His sheath, Medtronic, Inc., Minneapolis, MN) was used in the LBBAP group. In one study, SDES pacing lead (Solia S pacing lead, Biotronik SE & Co. KG, Berlin, Germany; 60 cm ventricular lead) was used with a success rate of 100%.

### 3.3 Quality assessment for included studies

In all observational studies the NOS was judged to be above 7 representing studies of a good quality (**Supplementary Table S1**). However, there were some concerns for the quality of RCTs (**Supplementary Figure S1**).

### 3.4 Pairwise meta-analysis

#### 3.4.1 Clinical outcomes

##### 3.4.1.1 Heart Failure Hospitalizations

Heart failure hospitalizations was assessed in 8 studies including a total of 1966 patients. LBBAP was associated with a lower risk of HFH (RR: 0.33, CI 95%: 0.21 to 0.50; I^2^=0%; *p* < 0.001; **Figure 2A**).

**Figure 2.**
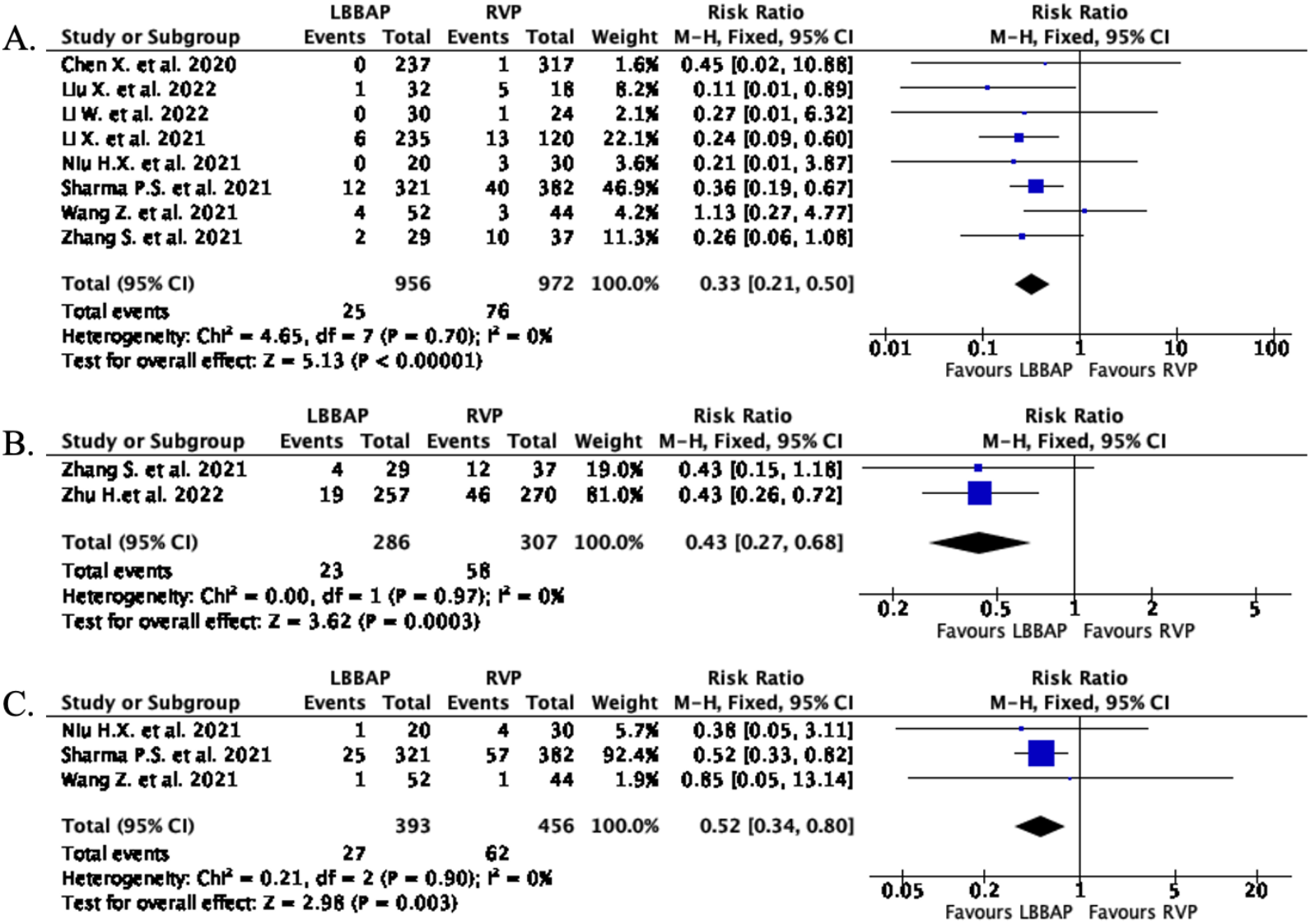
Forest plots of (A) HFH for LBBAP vs RVP group; (B) AF occurrence for LBBAP vs RVP group; (C) all-cause mortality for LBBAP vs RVP group. AF = atrial fibrillation, HFH = heart failure hospitalizations, LBBAP = left bundle branch area pacing, RVP = right ventricular pacing.

##### 3.4.1.2 Atrial Fibrillation occurrence

The occurrence of AF was assessed in two studies including 593 patients. In LBBAP group 23 total events of AF were reported versus 58 in the RVP group. LBBAP was associated with a lower risk of AF occurrence compared with RVP (RR: 0.43 CI 95%: 0.27 to 0.68; I^2^=0%; *p*<0.001, **Figure 2B**).

##### 3.4.1.3 All-cause mortality

All–cause mortality was assessed in 3 studies, including 849 patients, that reported 27 deaths in LBBAP group versus 62 in the RVP group. LBBAP was associated with a lower risk of all-cause mortality compared with RVP (RR:0.52 CI 95%:0.34 to 0.80; I^2^=0%; *p*=0.003, **Figure 2C**). These mortality events occurred during a mean follow-up period of 16.11 ± 8.41 months after implantation.

#### 3.4.2 Complications

Complications mainly referred to lead related complications and were reported in 20 studies. The rate of late lead perforation between the two groups was similar (0.18%, LBBAP vs 0.06%, RVP; *p*=0.350), whereas periprocedural lead perforation was higher in LBBAP group (0.77%, LBBAP vs 0%, RVP; *p*=0.020). On the contrary, the rate of lead dislodgement (into the right ventricle) was higher in the RVP group (0.65%, LBBAP vs 1.37%, RVP; *p*=0.040). However, the rate of lead related events in total had no difference between the two groups (1.67%, LBBAP vs 1.54%, RVP; *p*=0.780) (**Figure 3**).

**Figure 3.**
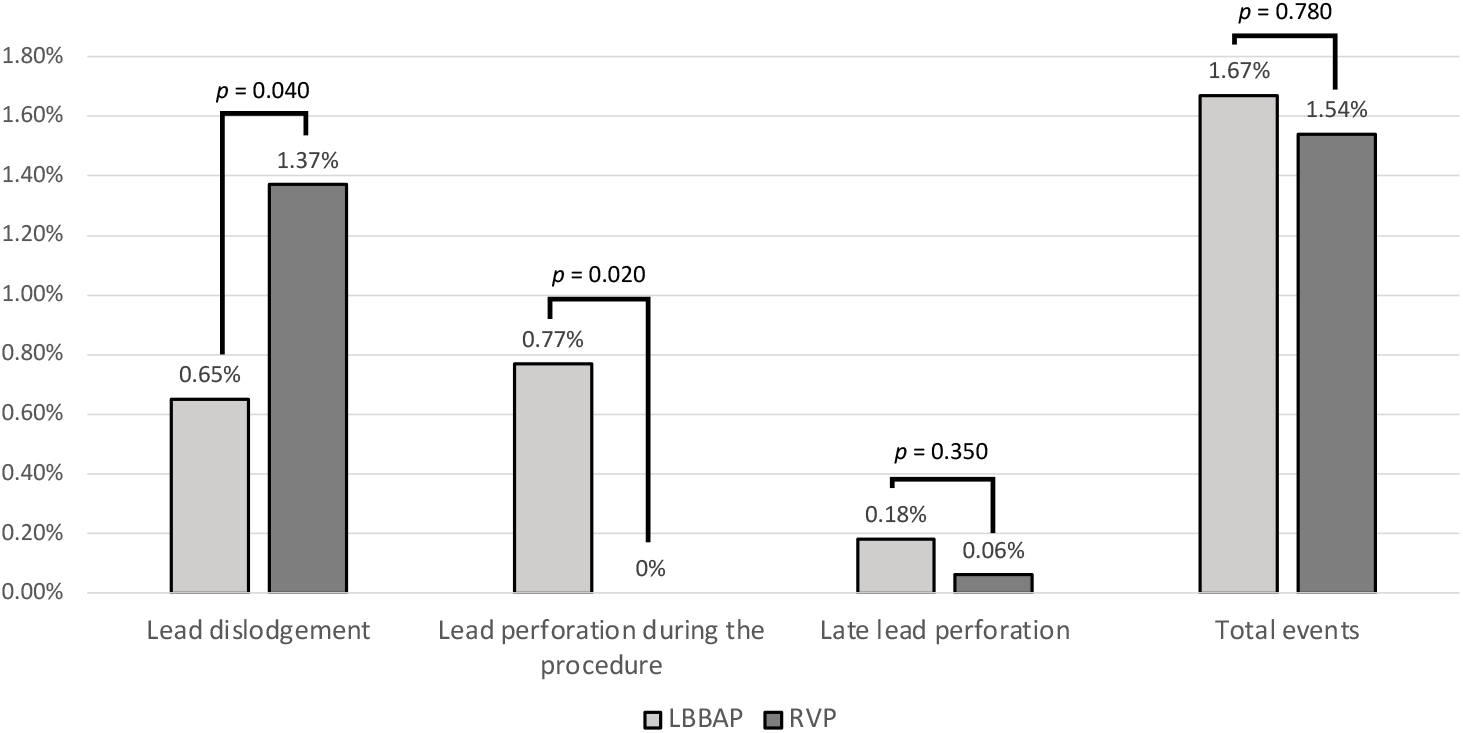
Lead related complications diagram for LBBAP vs RVP group. LBBAP = left bundle branch area pacing, RVP = right ventricular pacing.

#### 3.4.3 Ventricular electrical synchrony

##### 3.4.3.1 QRS duration

The QRSd at baseline was recorded in 23 studies. LBBAP preserved the baseline QRS (WMD: -1.92 msec, CI 95%: -6.03 to 2.19; I^2^=89%; *p*=0.360; **Figure 4A**). These results remained robust in the leave-one-out sensitivity analysis.

**Figure 4.**
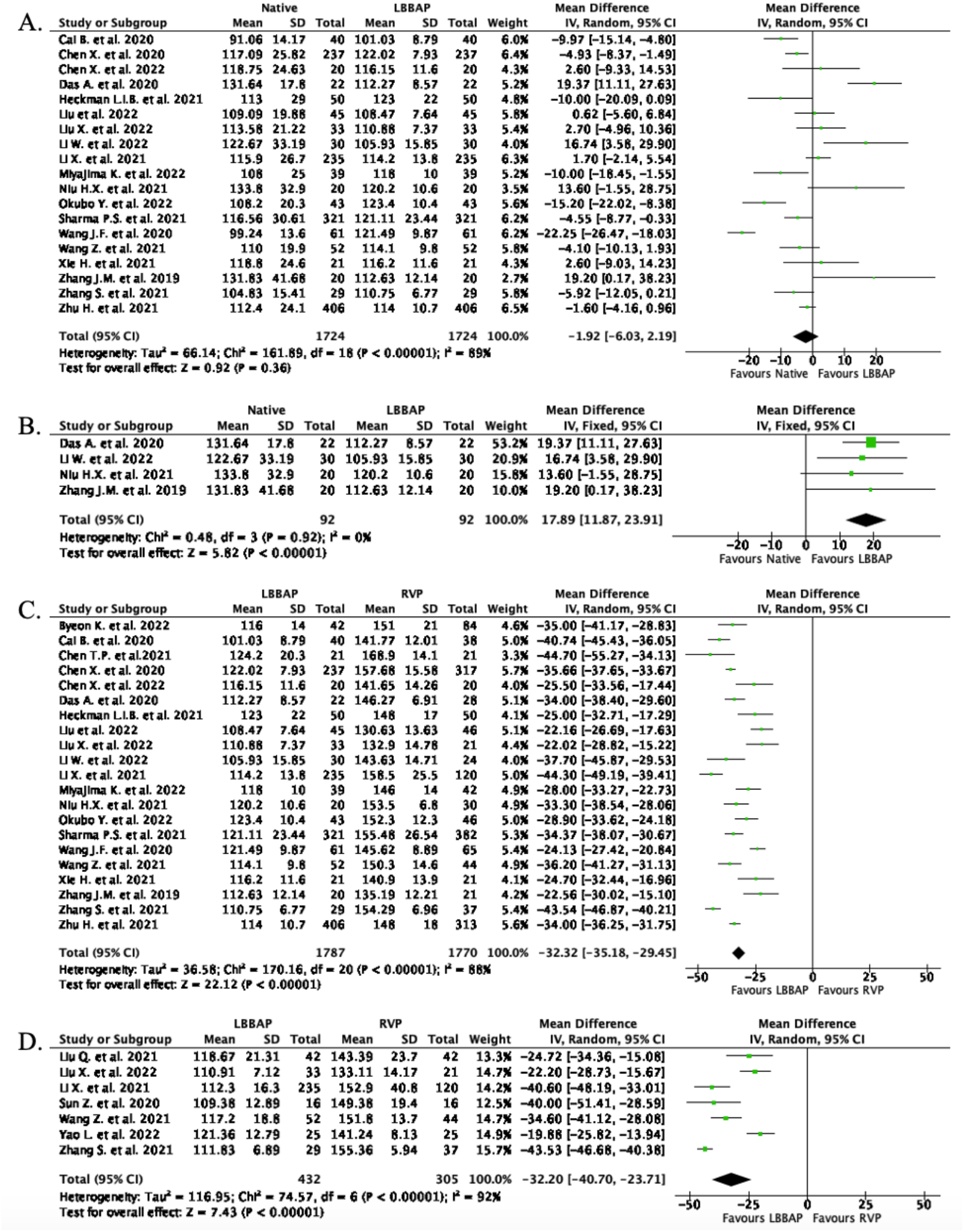
Forest plots of QRSd. (A) for native vs LBBAP group; (B) for native vs LBBAP group only in studies with baseline QRSd >120 ms; (C) for LBBAP vs RVP group following implantation;(D) for LBBAP vs RVP group at follow up. LBBAP = left bundle branch area pacing, QRSd = QRS duration, RVP = right ventricular pacing.

In the 4 trials where the baseline QRSd was longer than 120 msec, there was a QRSd shortening after LBBAP implantation (WMD: 17.89 msec, CI 95%: 11.87 to 23.91; I^2^=0%; *p*<0.001; **Figure 4B**). The QRSd was shorter immediately after LBBAP implantation (WMD: -32.32 msec, CI 95%: -35.18 to -29.45; I^2^=88%; *p*<0.001; **Figure 4C**) and at follow-up (WMD: -32.20 msec, CI 95%: -40.70 to -23.71; I^2^=92%; *p*<0.001; **Figure 4D**) compared with RVP, representing better ventricular electrical synchrony. These results remained robust in the leave-one-out sensitivity analysis.

##### 3.4.3.2 Stim-LVAT

Stim-LVAT was assessed in seven studies. Stim-LVAT in LBBAP group was shorter compared with RVP (WMD: -24.40 msec, CI 95%: -36.32 to -12.48; I^2^=98%; *p*<0.001; **supplementary fig. S2**). Sensitivity analysis found the results to be stable.

#### 3.4.4 LV mechanical synchrony

##### 3.4.4.1 Intraventricular mechanical synchrony

Intraventricular mechanical synchrony between native conduction and LBBAP immediately post implantation or at follow-up had no significant difference (SMD: 0.06, CI 95%: -0.11 to 0.24; I^2^=33%; *p*=0.480; **Figure 5A**).

**Figure 5.**
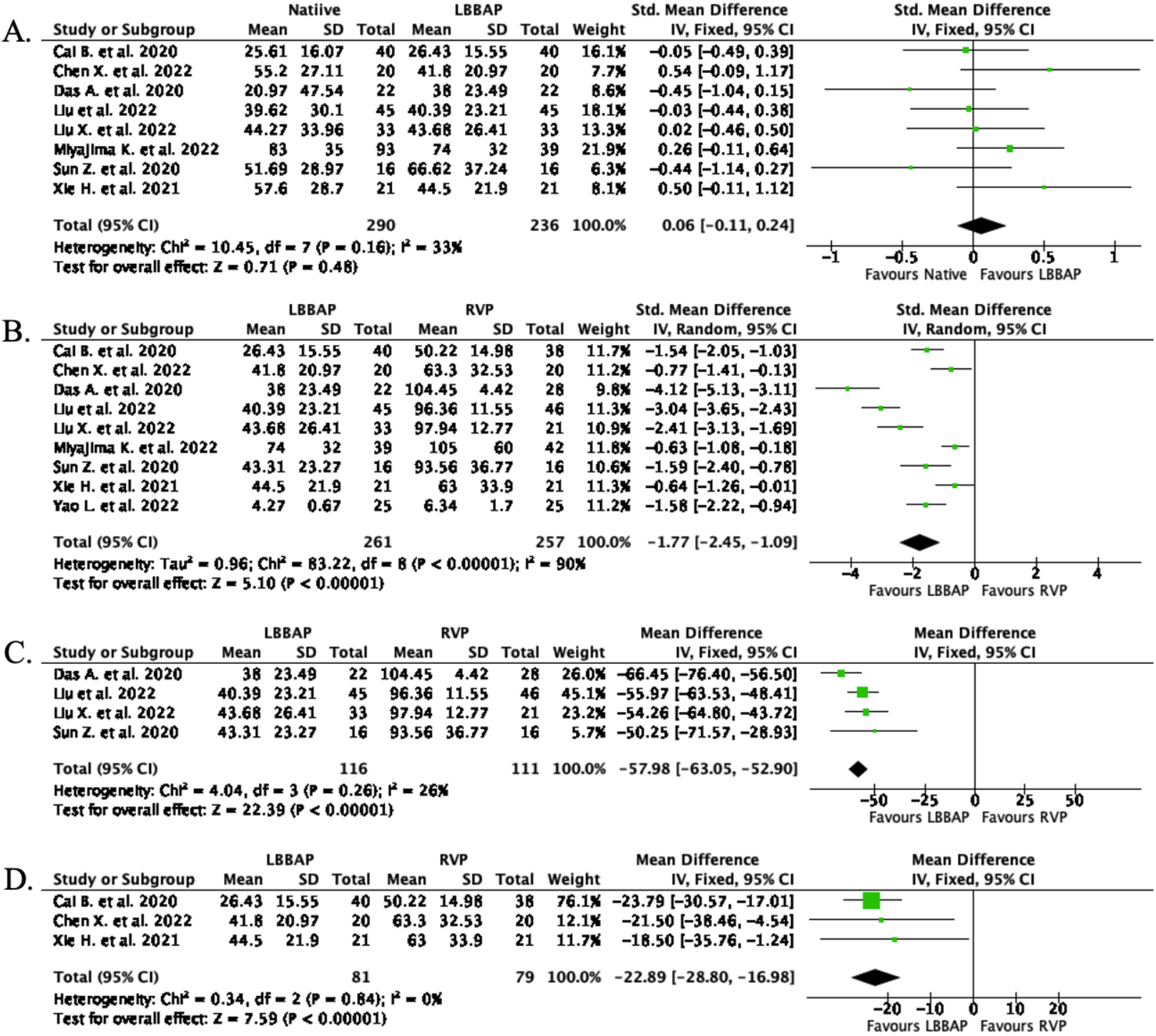
Forest plots of intraventricular mechanical synchrony. (A) for native vs LBBAP group; (B) for LBBAP vs RVP group immediately post implantation and at follow up; (C) for LBBAP vs RVP group using SPWMD; (D) for LBBAP vs RVP group using TS-12-SD. LBBAP = left bundle branch area pacing, RVP = right ventricular pacing, SPWMD = septal to posterior wall motion delay.

LBBAP had better intraventricular mechanical synchrony compared to RVP post implantation and at follow-up (SMD: -1.77, CI 95%: -2.45 to -1.09; I^2^=90 %; *p*<0.001; **Figure 5B**). Considering the high heterogeneity of the methods used to assess intraventricular synchrony, we performed a subgroup analysis including only studies that used the same way of measurement of intraventricular synchrony. Studies that measured intraventricular dyssynchrony with the SPWMD method, showed improved intraventricular synchrony with LBBAP (WMD: -57.98 msec, CI 95%: -63.05 to -52.90; I^2^=26%; *p*<0.001; **Figure 5C**). Similar, studies that used the TS-12-SD method showed improved synchrony with LBBAP compared to RVP (WMD: -22.89 msec, CI 95%: -28.80 to -16.98; I^2^= 0%; *p*<0.001; **Figure 5D**).

##### 3.4.4.2 Interventricular mechanical synchrony

We included studies that evaluated the presence of dyssynchrony between the two ventricles, using the interventricular mechanical delay (IVMD). However, each study implemented different methods for this purpose. The analysis showed that LBBAP ensured significantly better interventricular mechanical synchrony than RVP (SMD: -2.04, CI 95%: -2.32 to -1.76; I^2^=21%; *p*<0.001; **supplementary fig. S3**).

#### 3.4.5 LV systolic function

Left ventricular systolic function was assessed with LVEF which was reported at baseline and at follow-up in 11 studies and with left ventricular end diastolic diameter (LVEDD). There were no significant differences in LVEF and LVEDD at baseline between the two groups.

##### 3.4.5.1 Left ventricular ejection function

LBBAP was associated with a higher LVEF compared with RVP at follow-up (WMD: 2.89 %, CI 95%: 1.70 to 4.07; I2=56%; *p*<0.001; **Figure 6**). Sensitivity analysis showed consistency of the results. Of note a 2.89% difference in LVEF falls within the inter-observer variability of LVEF assessment and should thus be interpreted as the LVEF was similar in the two groups.

**Figure 6.**
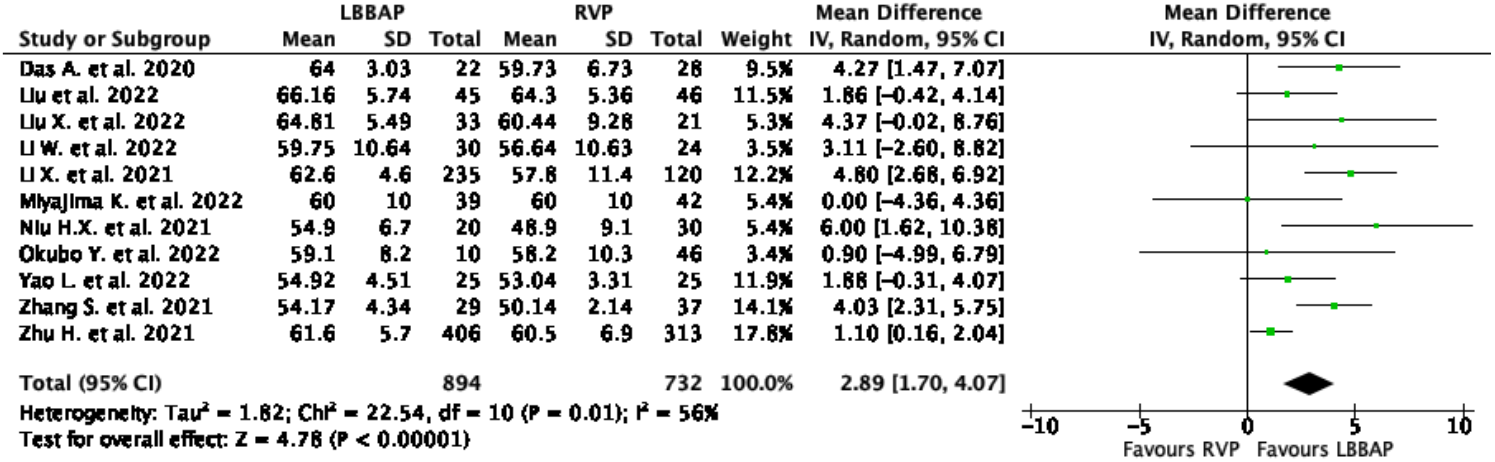
Forest plot of LVEF for LBBAP vs RVP group at follow-up. LVEF = left ventricle ejection fraction, LBBAP = left bundle branch area pacing, RVP = right ventricular pacing.

##### 3.4.5.2 Left ventricular end-diastolic diameter

Left ventricular end-diastolic diameter improved after LBBAP compared to native conduction (WMD: 1.82 mm, CI 95%: 1.14 to 2.50; I^2^=26%; *p*<0.001; **supplementary fig. S4A**). LVEDD was also marginally smaller in LBBAP group compared to RVP group (WMD: -2.74 mm, CI 95%: -4.41 to -1.07; I^2^= 73%; *p* = 0.001; **supplementary fig. S4B**). Sensitivity analysis showed that the results were stable. In both comparisons the differences cannot be considered significant from a clinical point of view.

#### 3.4.6 Pacing parameters

##### 3.4.6.1 Pacing threshold

Pacing threshold was assessed in 21 studies. The analysis showed that in LBBAP group, the pacing threshold immediately post implantation was low and similar with RVP group (WMD: 0.00 V/0.4 ms, CI 95%: -0.04 to 0.05; I^2^=88%; *p*=0.860; **supplementary fig. S5**). These results remained robust in the leave-one-out sensitivity analysis.

##### 3.4.6.2 Lead impedance

Lead impendence between LBBAP and RVP immediately post implantation was similar (WMD: -7.98 Ω, CI 95%: -25.78 to 9.83; I^2^=61%; *p* = 0.380; **supplementary fig. S6A**). Lead impedance in LBBAP was lower at follow-up than post implantation (WMD: 125.42 Ω, CI 95%: 85.01 to 165.83; I^2^=93%; *p*<0.001; **supplementary fig. S6B**), but not different than that of the RVP group (WMD: -19.10 Ω, CI 95%: -55.25 to 17.05; I^2^=93%; *p*=0.300; **supplementary fig. S6C**). Sensitivity analysis showed stability of the results.

##### 3.4.6.3 R wave amplitude

The R wave amplitude was compared in 17 studies. LBBAP achieved marginally higher R wave amplitudes than RVP post implantation (WMD: 0.85 mV, CI 95%: 0.21 to 1.48; I^2^=71%; *p*=0.009; **supplementary fig. S7A**) that remained higher also at follow-up (WMD: 1.22 mV, CI 95%: 0.23 to 2.20; I^2^=91%; *p*=0.020; **supplementary fig. S7B**). Sensitivity analysis showed that the results were consistent.

#### 3.4.7 Procedural Characteristics

Procedural duration in LBBAP group was significantly longer than in RVP group (WMD: 22.44 min, CI 95%: 11.53 to 33.36; I^2^=98%; *p*<0.001; **supplementary fig. S8A**). Similar, fluoroscopy time in LBBAP group was longer (WMD: 5.84 min, CI 95%: 3.49 to 8.20; I^2^=98%; *p*<0.001; **supplementary fig. S8B**).

### 3.5 Publication bias

Funnel plot asymmetry and *p* value for Egger’s test < 0.05 was only detected for the outcomes of QRSd native vs LBBAP and intraventricular mechanical synchrony LBBAP vs RVP. For all the other outcomes publication bias was not detected. Specifically for the outcome of QRSd, studies that report a significant shortening of the QRSd from baseline with low accuracy are missing. This may not represent true publication bias, but rather the fact that in most studies the baseline QRSd was normal and thus LBBAP didn’t shorten QRSd beyond baseline. For the outcome of intraventricular mechanical synchrony, studies reporting large magnitude of worsening intraventricular dyssynchrony with low accuracy are missing from the literature likely representing true publication bias.

## 4. Discussion

In this meta-analysis we demonstrate that LBBAP compared with RVP for bradycardia and conduction system disorders is associated with improved clinical outcomes. Our most important findings are the following: a) LBBAP is associated with less HFH, lower AF occurrence rates and a reduction in all-cause mortality; b) Lead related complications are similar in both groups; c) LBBAP preserves the baseline QRSd if shorter than 120 ms and is related with narrower QRS than RVP both immediately post implantation and at FU; d) LBBAP results in more synchronized mechanical contraction of the ventricles than RVP by achieving better intraventricular and interventricular synchronization; e) LBBAP achieves low and stable pacing thresholds like that of RVP and lead parameters at follow-up are similar to those of RVP

Over the last years there has been an increasing utilization of LBBAP as an alternative pacing modality. The two clinical conditions where LBBAP is being utilized is (a) in patients with an indication for Cardiac Resynchronization Therapy (CRT), and (b) in patients with bradycardia and conduction system disorders. The use of LBBAP as a first-line pacing modality for the indication of bradycardia or conduction system disease is currently not supported by the guidelines. In this Systematic Review and Meta-analysis, we explored the potential benefits of LBBAP in patients with bradycardia or conduction system disease.

Due to the lack of randomized clinical studies, we cannot support so far LBBAP as an upfront strategy for all comers. With regards to patients with bradycardia and conduction system disorders two large, multicenter, RCTs are ongoing. The LEAP-Block (NCT04730921)^24^ and the PROTECT-SYNC (NCT05585411) ^25^ trials will compare LBBAP with RVP with the outcomes all-cause mortality, HFH and rates of need for device upgrade due to development heart failure. An ongoing randomized trial by our group (NCT05129098)^26^ examines improvement of mechanical dyssynchrony measured by advanced echocardiographic parameters in LBBAP vs RVP patients.

With regards to patients with an indication for CRT there are a few observational studies available^27-30^ that show the positive effects of LBBAP compared to biventricular pacing (BVP) but only one RCT (LBBP-RESYNC Trial)^31^ has already been completed to date. According to this RCT, LBBAP achieves better LVEF improvement than BVP in heart failure patients with nonischemic cardiomyopathy and LBBB. In a different trial^32^ conducted by the LBBAP Collaborative Study Group LBBAP was successfully used as a bail out strategy for BVP in patients that present difficulties in implantation of the Coronary Sinus (CS) lead, or who are nonresponders to BVP.

We aimed to summarize studies who examine LBBAP as an alternative mode of pacing in patients with bradycardia and conduction system disorders. The effects of LBBAP in the field of CRT was outside the scope of this meta-analysis. For this, we included only studies in patients with LVEF >35%. In all the included studies, the mean LVEF of the participants was >50%. As such, we didn’t have discrete evidence showing that patients with LVEF=35-50% would benefit more from LBBAP versus RVP. In the BLOCK HF trial^33^ that included patients with atrioventricular nodal (AVN) disease and an indication for pacemaker and LVEF 35-50%, it was found that BVP is more beneficial than conventional RVP alone. For this, in patients with EF <40% who are expected to have a significant burden of pacing, BVP is recommended by most recent guidelines^2^ with a class IA indication. However, from our analysis we cannot indicate that LBBAP is more beneficial than RVP in this subgroup of patients as we didn’t have discrete data from this subgroup.

The present study showed that LBBAP is associated with better clinical outcomes, including lower HFH rates. However, we didn’t observe clinically significant differences in LVEF at follow up between the two groups (WMD: 2.89%). A potential explanation is that in patients with RVP, the asynchronous contraction of the lateral wall of the left ventricle causes mitral regurgitation. As a result, some blood volume regurgitates in the left atrium during systole. The LVEF appears to be normal, because the end systolic volume remains the same although a proportion of blood volume goes backwards, increasing this way the risk of heart failure. As shown in our analysis, LBBAP achieves better mechanical synchrony of the ventricles, and it is thus possible that the favorable clinical outcomes are mediated via restoration of mechanical synchrony rather than preservation of LV ejection fraction. Furthermore, it is possible that some of our patients had some degree of diastolic dysfunction (DD) at baseline. According to Jeong et al,^34^ RVP increased the risk for HF in patients with DD at baseline, even if their systolic LV function was preserved.

An important number of patients in the included studies had BBB. It was found that LBBAP corrected BBB in 84.2% of the cases. Failure to BBB correction could be due to the fact that in a small proportion of patients the region of the block was localized distal to the LBB. Thus, criteria need to be developed to guide patient selection who could benefit from corrective LBBAP as 36% of the patients with LBBB have conduction block distal to the LBB.^35^

## 5. Limitations

First, most of the included studies were observational studies. Only four were RCTs and not sufficiently powered for all outcomes. Thus, we pooled the data from RCTs with that from observational studies, which may introduce some uncontrolled bias. Second, many of the studies had a small sample size and short follow up period which may lead to underestimation of the actual results. Third, there were studies that had also an HBP group, and some trials had a crossover design which may introduce some bias to the results. Four, despite inevitably introducing some heterogeneity, studies were included regardless of the follow-up time to capture the full view of pacing modalities. Also, a high heterogeneity was present in some of the outcomes. For this a random-effects model was used and sensitivity analysis was performed in each case and indicated consistency and stability of the results. Five, differences in methodological quality of each study, publication bias and the exclusion of studies that didn’t report the outcomes we needed, may contribute to the introduction of bias in our results.

## 6. Conclusions

LBBAP is associated with significantly lower HFH, all-cause mortality, AF occurrence rates, and similar lead related complication rates in comparison to RVP in patients with bradycardia and conduction system disorders. It achieves better electromechanical ventricular synchrony, low, and stable pacing thresholds, and high R-waves. However, larger randomized studies are needed to extract robust data and further verify the results about the outcomes and the safety of this method, as presented in our meta-analysis.

## Data Availability

Data are available upon reasonable request from the corresponding author at.

## Non-standard Abbreviations and Acronyms

AF: atrial fibrillation
AVB: atrioventricular block
AVN: atrioventricular nodal
BBB: bundle branch block
BVP: biventricular pacing
CRT: cardiac resynchronization therapy
DD: diastolic dysfunction
FU: follow-up
HBP: His-bundle pacing
HFH: heart failure hospitalization
IVMD: interventricular mechanical delay
LBB: left bundle branch
LBBAP: left bundle branch area pacing
LVEDD: left ventricular end-diastolic diameter
LVEF: left ventricular ejection fraction
RCT: randomized controlled trial
RVP: right ventricular pacing
SND: sinus node dysfunction
SPWMD: septal to posterior wall motion delay
Stim-LVAT: stim to left ventricular activation time

## Acknowledgments

None

## Sources of Funding

None

## Disclosures

GL has received honoraria for speaking at educational events from Medtronic. KNA has received funds from Abbott and Haemonetics. These are not relevant to the current publication or left bundle branch pacing. KNA has no conflicts of interest to disclose. CPG reports research grant support and/or speaker/consulting honoraria from: Abbott Diabetes, AINexus, Amgen, AstraZeneca, Bayer, Boehringer Ingelheim, Bristol Myers Squibb, British Heart Foundation, Cardiomatics, Chiesi, GPRI Research B.V., Daiichi-Sankyo, Eli Lilly, European Society of Cardiology, Horizon2020, iRhythm, Medisetter, Menarini, National Institute for Health Research, Novartis, Organon, Pfizer, Raisio Group, Vifor Pharma, Wondr medical, Zydus. These are not relevant to the current publication or left bundle branch pacing.

## Supplemental Material

Supplemental Methods

Table S1

Figure S1-S9

## Data availability statement

Data are available upon reasonable request from the corresponding author at.

---

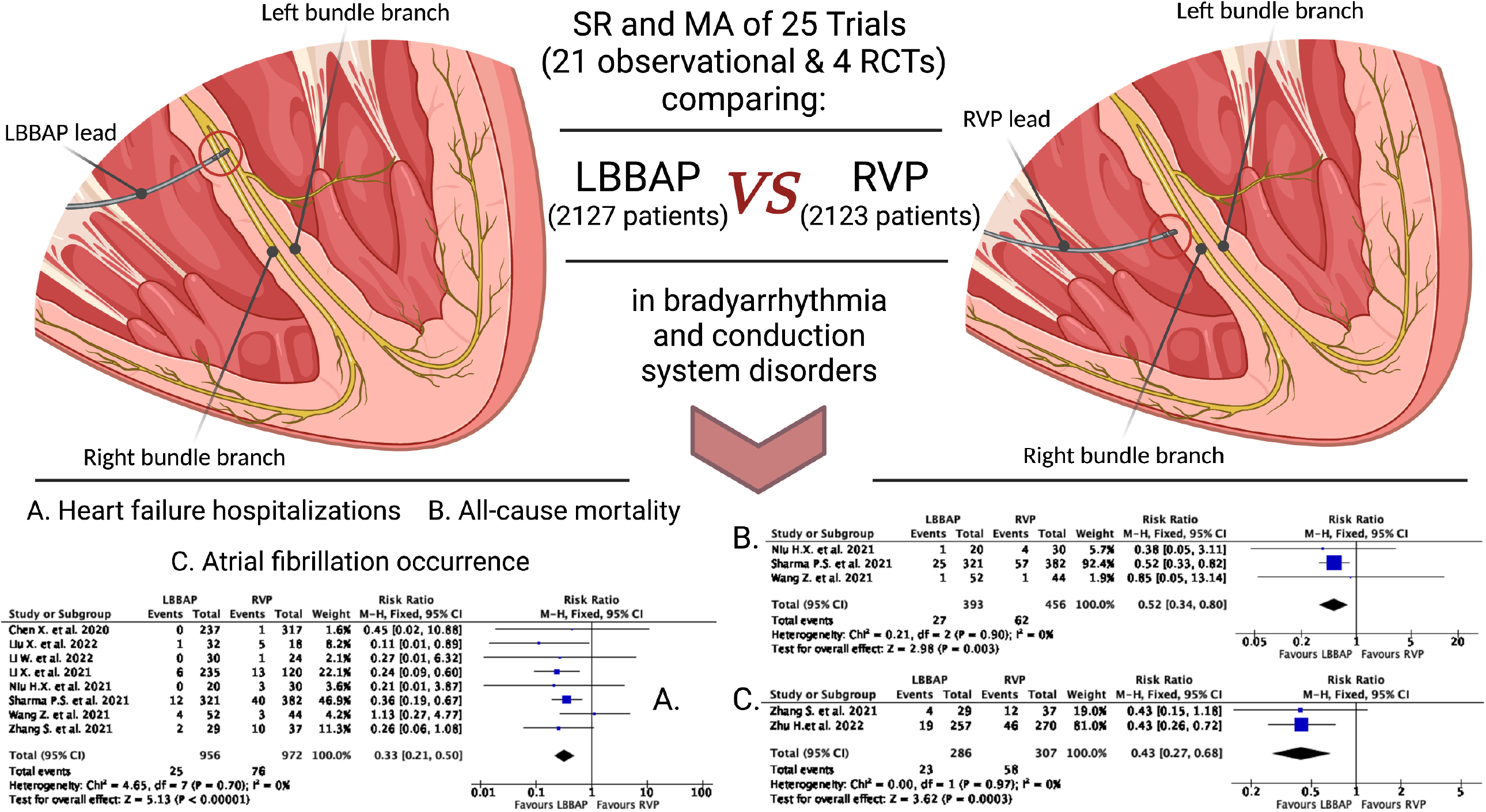

## Notes

### Competing Interest Statement

The authors have declared no competing interest.

