## Supplement for "Safety and efficacy of left bundle branch area pacing compared with right ventricular pacing in patients with bradyarrhythmia and conduction system disorders: a systematic review and meta-analysis"

### Supplementary material

#### 1. Prisma checklist

| Section and Topic | Item # | Checklist item | Location where item is reported |
|---|---|---|---|
| **TITLE** | | | |
| Title | 1 | Identify the report as a systematic review. | p.1 |
| **ABSTRACT** | | | |
| Abstract | 2 | See the PRISMA 2020 for Abstracts checklist. | p. 2 |
| **INTRODUCTION** | | | |
| Rationale | 3 | Describe the rationale for the review in the context of existing knowledge. | p.5 |
| Objectives | 4 | Provide an explicit statement of the objective(s) or question(s) the review addresses. | p.5,6 |
| **METHODS** | | | |
| Eligibility criteria | 5 | Specify the inclusion and exclusion criteria for the review and how studies were grouped for the syntheses. | p.6,7 |
| Information sources | 6 | Specify all databases, registers, websites, organisations, reference lists and other sources searched or consulted to identify studies. Specify the date when each source was last searched or consulted. | p.6 |
| Search strategy | 7 | Present the full search strategies for all databases, registers and websites, including any filters and limits used. | supplement |
| Selection process | 8 | Specify the methods used to decide whether a study met the inclusion criteria of the review, including how many reviewers screened each record and each report retrieved, whether they worked independently, and if applicable, details of automation tools used in the process. | p.6,7 |
| Data collection process | 9 | Specify the methods used to collect data from reports, including how many reviewers collected data from each report, whether they worked independently, any processes for obtaining or confirming data from study investigators, and if applicable, details of automation tools used in the process. | p.8 |
| Data items | 10a | List and define all outcomes for which data were sought. Specify whether all results that were compatible with each outcome domain in each study were sought (e.g. for all measures, time points, analyses), and if not, the methods used to decide which results to collect. | p.7,8 |
| | 10b | List and define all other variables for which data were sought (e.g. participant and intervention characteristics, funding sources). Describe any assumptions made about any missing or unclear information. | p.8 |
| Study risk of bias assessment | 11 | Specify the methods used to assess risk of bias in the included studies, including details of the tool(s) used, how many reviewers assessed each study and whether they worked independently, and if applicable, details of automation tools used in the process. | p.9 |
| Effect measures | 12 | Specify for each outcome the effect measure(s) (e.g. risk ratio, mean difference) used in the synthesis or presentation of results. | p.9,10 |
| Synthesis methods | 13a | Describe the processes used to decide which studies were eligible for each synthesis (e.g. tabulating the study intervention characteristics and comparing against the planned groups for each synthesis (item #5)). | p.8 |
| | 13b | Describe any methods required to prepare the data for presentation or synthesis, such as handling of missing summary statistics, or data conversions. | p.9 |

| Section and Topic | Item # | Checklist item | Location where item is reported |
|---|---|---|---|
| | 13c | Describe any methods used to tabulate or visually display results of individual studies and syntheses. | p.8,9 |
| | 13d | Describe any methods used to synthesize results and provide a rationale for the choice(s). If meta-analysis was performed, describe the model(s), method(s) to identify the presence and extent of statistical heterogeneity, and software package(s) used. | p.9,10 |
| | 13e | Describe any methods used to explore possible causes of heterogeneity among study results (e.g. subgroup analysis, meta-regression). | p.9,10 |
| | 13f | Describe any sensitivity analyses conducted to assess robustness of the synthesized results. | p.10 |
| Reporting bias assessment | 14 | Describe any methods used to assess risk of bias due to missing results in a synthesis (arising from reporting biases). | p.10 |
| Certainty assessment | 15 | Describe any methods used to assess certainty (or confidence) in the body of evidence for an outcome. | p.9,10 |
| **RESULTS** | | | |
| Study selection | 16a | Describe the results of the search and selection process, from the number of records identified in the search to the number of studies included in the review, ideally using a flow diagram. | p.10 |
| | 16b | Cite studies that might appear to meet the inclusion criteria, but which were excluded, and explain why they were excluded. | - |
| Study characteristics | 17 | Cite each included study and present its characteristics. | Table 1 |
| Risk of bias in studies | 18 | Present assessments of risk of bias for each included study. | supplement |
| Results of individual studies | 19 | For all outcomes, present, for each study: (a) summary statistics for each group (where appropriate) and (b) an effect estimate and its precision (e.g. confidence/credible interval), ideally using structured tables or plots. | p.21 |
| Results of syntheses | 20a | For each synthesis, briefly summarise the characteristics and risk of bias among contributing studies. | p.11-16 |
| | 20b | Present results of all statistical syntheses conducted. If meta-analysis was done, present for each the summary estimate and its precision (e.g. confidence/credible interval) and measures of statistical heterogeneity. If comparing groups, describe the direction of the effect. | p.11-16 |
| | 20c | Present results of all investigations of possible causes of heterogeneity among study results. | p.11-16 |
| | 20d | Present results of all sensitivity analyses conducted to assess the robustness of the synthesized results. | p.11-16 |
| Reporting biases | 21 | Present assessments of risk of bias due to missing results (arising from reporting biases) for each synthesis assessed. | p.16,17 |
| Certainty of evidence | 22 | Present assessments of certainty (or confidence) in the body of evidence for each outcome assessed. | p.11-16 |
| **DISCUSSION** | | | |
| Discussion | 23a | Provide a general interpretation of the results in the context of other evidence. | p.16-19 |
| | 23b | Discuss any limitations of the evidence included in the review. | p.19 |
| | 23c | Discuss any limitations of the review processes used. | p.19 |
| | 23d | Discuss implications of the results for practice, policy, and future research. | p.16-19 |
| **OTHER INFORMATION** | | | |
| Registration and | 24a | Provide registration information for the review, including register name and registration number, or state that the review was not | p.5 |

| Section and Topic | Item # | Checklist item | Location where item is reported |
|---|---|---|---|
| protocol | | registered. | |
| | 24b | Indicate where the review protocol can be accessed, or state that a protocol was not prepared. | p.5 |
| | 24c | Describe and explain any amendments to information provided at registration or in the protocol. | p.5 |
| Support | 25 | Describe sources of financial or non-financial support for the review, and the role of the funders or sponsors in the review. | p.21 |
| Competing interests | 26 | Declare any competing interests of review authors. | p.21 |
| Availability of data, code and other materials | 27 | Report which of the following are publicly available and where they can be found: template data collection forms; data extracted from included studies; data used for all analyses; analytic code; any other materials used in the review. | p.21 |

#### 2. Formulation of research question using PICOT (Population, Intervention, Comparison, Outcomes, Type of studies) question frame

The research question was structured using the PICOT question frame as follows:

**Population:** Patients with bradyarrhythmia and conduction system disorders.

**Intervention:** Left bundle branch area pacing

**Comparison:** Right ventricular pacing

**Outcomes:**

a) Clinical outcomes, b) Lead related complications, c) Ventricular electrical synchrony, assessed by paced QRS duration, and stim to LVAT (left ventricle activation time), d) Left ventricular mechanical synchrony, including intraventricular and interventricular synchrony, e) Left ventricular systolic function assessed by left ventricle ejection fraction (LVEF) and left ventricle end diastolic diameter (LVEDD), f) Pacing parameters including pacing threshold, ventricular impedance, R wave amplitude, g) Procedural characteristics including procedural duration, fluoroscopy time, procedural successs rate, probability of recording LBB potential and correction of BBB.

**Type of included Studies:**

Randomized control trials (RCTs) and observational studies.

3. **Search Terms and search strategy with full results**

Database: **Ovid MEDLINE(R) ALL** 1946 to November 10, 2022

Search strategy:

| # | Searches | Results |
|---|---|---|
| 1 | LBBP.mp | 142 |
| 2 | Left bundle branch pacing.mp | 279 |
| 3 | left bundle branch area pacing.mp | 153 |
| 4 | LBBaP.mp | 105 |
| 5 | LBB*a*P.mp | 105 |
| 6 | 1 or 2 or 3 or 4 or 5 | 410 |
| 7 | RVP.mp. | 749 |
| 8 | RV Pacing.mp. | 637 |
| 9 | Right Ventricular Pacing.mp. | 1425 |
| 10 | Right Ventricular Septal.mp. | 143 |
| 11 | Right Ventricular Apical.mp. | 444 |
| 12 | 7 or 8 or 9 or 10 or 11 | 2944 |
| 13 | 6 and 12 | 94 |

Database(s): **Embase Classic + Embase** 1947 to November 10, 2022

Search Strategy:

| # | Searches | Results |
|---|---|---|
| 1 | LBBP.mp | 214 |
| 2 | Left bundle branch pacing.mp | 385 |
| 3 | left bundle branch area pacing.mp | 230 |
| 4 | LBBaP.mp | 161 |
| 5 | LBB*a*P.mp | 161 |
| 6 | 1 or 2 or 3 or 4 or 5 | 594 |
| 7 | RVP.mp. | 1606 |
| 8 | RV Pacing.mp. | 1714 |
| 9 | Right Ventricular Pacing.mp. | 2441 |
| 10 | Right Ventricular Septal.mp. | 258 |
| 11 | Right Ventricular Apical.mp. | 765 |
| 12 | 7 or 8 or 9 or 10 or 11 | 5705 |
| 13 | 6 and 12 | 128 |

Database(s): **PubMed** until November 10, 2022

Search Strategy:

((lbbp) OR (left bundle branch pacing)) AND ((right ventricular pacing) OR (rv pacing) OR (rvp))

Results: 1096

**Supplementary Table S1: Quality assessment of the included observational studies according to the Newcastle-Ottawa Scale.**

| Study ID | Selection | | | | Comparability | Outcome | | | Overall |
|---|---|---|---|---|---|---|---|---|---|
| | Representativeness of the exposed cohort | Selection of the non exposed cohort | Ascertainment of exposure | Demonstration that outcome of interest was not present at start of study | Comparability of cohorts on the basis of the design or analysis | Assessment of outcome | Was follow-up long enough for outcomes to occur | Adequacy of follow up of cohorts | |
| Byeon et al. 2022 | * | * | * | * | ** | * | * | * | **9** |
| Cai et al. 2020 | * | * | * | * | ** | * | * | * | **9** |
| Chen X. et al. 2022 | * | * | * | * | ** | * | * | | **8** |
| Chen X. et al. 2020 | * | * | * | * | * | * | * | | **7** |
| Li et al. 2021 | * | * | * | * | ** | * | * | * | **9** |
| Li et al. 2022 | * | * | * | * | ** | * | * | * | **9** |
| Liu et al. 2022 | * | * | * | * | ** | * | * | * | **9** |
| Liu X. et al. 2022 | * | * | * | * | ** | * | | * | **8** |
| Miyajima et al. 2022 | * | * | * | * | * | * | * | * | **8** |
| Niu et al. 2021 | * | * | * | * | ** | * | * | * | **9** |
| Sharma et al. 2021 | * | * | * | * | ** | * | * | * | **9** |
| Sun et al. 2020 | * | * | * | * | ** | * | | * | **8** |
| Xie et al. 2021 | * | * | * | * | ** | * | | * | **8** |
| Zhang et al. 2019 | | * | * | * | * | * | * | * | **7** |
| Zhang et al. 2021 | * | * | * | * | ** | * | * | * | **9** |
| Zhu et al. 2021 | * | * | * | * | ** | * | * | * | **9** |
| Zhu et al. 2022 | * | * | * | * | * | * | * | * | **8** |
| Wang et al. 2021 | * | * | * | * | ** | * | * | * | **9** |
| Chen TP. et al.2021 | * | * | * | * | ** | * | * | * | **9** |
| Heckman et al. 2021 | * | * | * | * | ** | * | * | * | **9** |
| Okubo et al. 2022 | * | * | * | * | ** | * | * | * | **9** |

**Supplementary Figure S1: Quality assessment of the included RCTs according to the Cochrane Risk of Bias 2 assessment tool (ROB 2.0)**

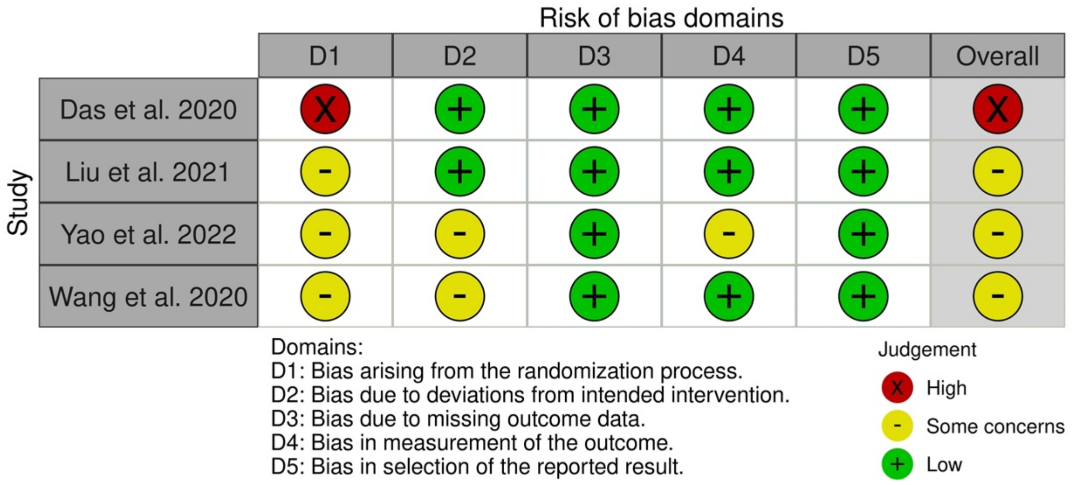


**Supplementary Figure S2:** Forest plot of stim-LVAT in LBBAP vs RVP group.

| | LBBAP | | | RVP | | | | Mean Difference |
|---|---|---|---|---|---|---|---|---|
| Study or Subgroup | Mean | SD | Total | Mean | SD | Total | Weight | IV, Random, 95% CI |
| Chen T.P. et al.2021 | 62.9 | 16.5 | 21 | 119.1 | 12.2 | 21 | 13.7% | −56.20 [−64.98, −47.42] |
| Chen X. et al. 2022 | 66.47 | 7.87 | 20 | 89.8 | 14.8 | 20 | 14.0% | −23.33 [−30.68, −15.98] |
| Heckman L.I.B. et al. 2021 | 78 | 11 | 50 | 105 | 11 | 50 | 14.5% | −27.00 [−31.31, −22.69] |
| Li W. et al. 2022 | 72.8 | 12.07 | 30 | 86.29 | 8.71 | 24 | 14.3% | −13.49 [−19.04, −7.94] |
| Miyajima K. et al. 2022 | 66 | 12 | 39 | 89 | 15 | 42 | 14.3% | −23.00 [−28.90, −17.10] |
| Okubo Y. et al. 2022 | 68.3 | 10 | 43 | 95.2 | 11.9 | 46 | 14.5% | −26.90 [−31.46, −22.34] |
| Zhu H. et al. 2021 | 73.9 | 13.4 | 406 | 76.7 | 15.4 | 313 | 14.7% | −2.80 [−4.95, −0.65] |
| Total (95% CI) | | | 609 | | | 516 | 100.0% | −24.40 [−36.32, −12.48] |

Heterogeneity: $Tau^2 = 250.24$; $Chi^2 = 274.04$, df = 6 ($P < 0.00001$); $I^2 = 98\%$
Test for overall effect: Z = 4.01 ($P < 0.0001$)

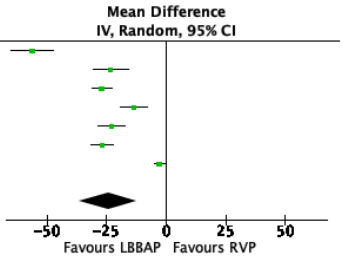


**Supplementary Figure S3:** Forest plot of interventricular mechanical synchrony for LBBAP vs RVP group.

| | LBBAP | | | RVP | | | | Std. Mean Difference |
|---|---|---|---|---|---|---|---|---|
| Study or Subgroup | Mean | SD | Total | Mean | SD | Total | Weight | IV, Fixed, 95% CI |
| Chen X. et al. 2022 | −19.25 | 18.43 | 20 | 22.85 | 22.05 | 20 | 13.1% | −2.03 [−2.81, −1.25] |
| Das A. et al. 2020 | 25.2 | 4.12 | 22 | 42.91 | 12.58 | 28 | 17.9% | −1.78 [−2.44, −1.11] |
| Liu et al. 2022 | 27.68 | 4.33 | 45 | 39.88 | 5.83 | 46 | 27.2% | −2.35 [−2.89, −1.81] |
| Liu X. et al. 2022 | 28.16 | 4.76 | 33 | 40.28 | 6.97 | 21 | 17.0% | −2.09 [−2.77, −1.41] |
| Sun Z. et al. 2020 | 10.23 | 3.07 | 16 | 39.94 | 14.81 | 16 | 8.1% | −2.71 [−3.70, −1.72] |
| Xie H. et al. 2021 | −14.9 | 28.3 | 21 | 22.6 | 21.4 | 21 | 16.7% | −1.47 [−2.16, −0.78] |
| Total (95% CI) | | | 157 | | | 152 | 100.0% | −2.04 [−2.32, −1.76] |

Heterogeneity: $Chi^2 = 6.32$, df = 5 ($P = 0.28$); $I^2 = 21\%$
Test for overall effect: Z = 14.22 ($P < 0.00001$)

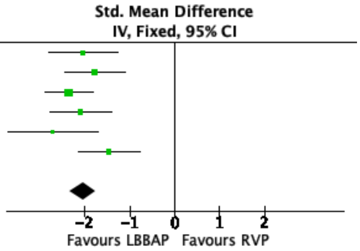

**Supplementary Figure S4.** Forest plots of LVEDD. (A)for native vs LBBAP group; (B) for LBBAP vs RVP group.

A.

| Study or Subgroup | Native Mean | Native SD | Native Total | LBBAP Mean | LBBAP SD | LBBAP Total | Weight | Mean Difference IV, Fixed, 95% CI |
|---|---|---|---|---|---|---|---|---|
| Liu et al. 2022 | 47.17 | 4.21 | 45 | 46.17 | 3.45 | 45 | 18.2% | 1.00 [−0.59, 2.59] |
| Liu X. et al. 2022 | 47.48 | 4.69 | 33 | 47.09 | 4.47 | 33 | 9.4% | 0.39 [−1.82, 2.60] |
| Li W. et al. 2022 | 50.73 | 6.54 | 30 | 47.39 | 6.95 | 30 | 3.9% | 3.34 [−0.07, 6.75] |
| Li X. et al. 2021 | 49.4 | 6.6 | 235 | 46.6 | 5.2 | 235 | 39.8% | 2.80 [1.73, 3.87] |
| Niu H.X. et al. 2021 | 52.7 | 8.1 | 20 | 49.7 | 5.6 | 20 | 2.5% | 3.00 [−1.32, 7.32] |
| Okubo Y. et al. 2022 | 48.71 | 3.27 | 29 | 47.58 | 3.29 | 29 | 16.1% | 1.13 [−0.56, 2.82] |
| Zhang S. et al. 2021 | 48.3 | 5.2 | 43 | 47.3 | 4.9 | 43 | 10.1% | 1.00 [−1.14, 3.14] |
| Total (95% CI) | | | 435 | | | 435 | 100.0% | 1.82 [1.14, 2.50] |

Heterogeneity: $Chi^2 = 8.08$, df = 6 (P = 0.23); $I^2 = 26\%$
Test for overall effect: Z = 5.27 (P < 0.00001)

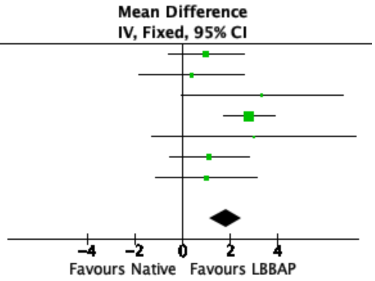


B.

| Study or Subgroup | LBBAP Mean | LBBAP SD | LBBAP Total | RVP Mean | RVP SD | RVP Total | Weight | Mean Difference IV, Random, 95% CI |
|---|---|---|---|---|---|---|---|---|
| Zhang S. et al. 2021 | 47.3 | 4.9 | 43 | 49.79 | 1.85 | 37 | 17.7% | −2.49 [−4.07, −0.91] |
| Okubo Y. et al. 2022 | 47.58 | 3.29 | 29 | 47.1 | 6.2 | 46 | 15.6% | 0.48 [−1.67, 2.63] |
| Niu H.X. et al. 2021 | 49.7 | 5.6 | 20 | 55 | 7.7 | 30 | 10.4% | −5.30 [−8.99, −1.61] |
| Li X. et al. 2021 | 46.6 | 5.2 | 235 | 51.7 | 7.5 | 120 | 18.0% | −5.10 [−6.60, −3.60] |
| Li W. et al. 2022 | 47.39 | 6.95 | 30 | 49.23 | 5.55 | 24 | 11.5% | −1.84 [−5.17, 1.49] |
| Liu X. et al. 2022 | 47.09 | 4.47 | 33 | 51.28 | 7.58 | 21 | 10.7% | −4.19 [−7.77, −0.61] |
| Liu et al. 2022 | 46.17 | 3.45 | 45 | 47.67 | 6.12 | 46 | 16.1% | −1.50 [−3.54, 0.54] |
| Total (95% CI) | | | 435 | | | 324 | 100.0% | −2.74 [−4.41, −1.07] |

Heterogeneity: $Tau^2 = 3.45$; $Chi^2 = 22.32$, df = 6 (P = 0.001); $I^2 = 73\%$
Test for overall effect: Z = 3.21 (P = 0.001)

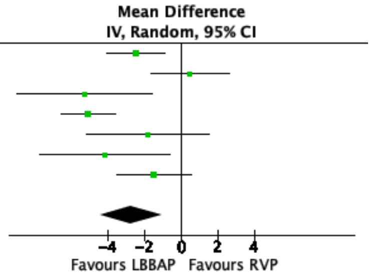


**Supplementary Figure S5.** Forest plot of pacing threshold for LBBAP vs RVP group at implantation.

| Study or Subgroup | LBBAP Mean | LBBAP SD | LBBAP Total | RVP Mean | RVP SD | RVP Total | Weight | Mean Difference IV, Random, 95% CI |
|---|---|---|---|---|---|---|---|---|
| Byeon K. et al. 2022 | 0.8 | 0.2 | 42 | 0.7 | 0.2 | 84 | 5.1% | 0.10 [0.03, 0.17] |
| Cai B. et al. 2020 | 0.49 | 0.22 | 40 | 0.62 | 0.18 | 38 | 4.8% | −0.13 [−0.22, −0.04] |
| Chen T.P. et al.2021 | 0.7 | 0.2 | 21 | 0.8 | 0.2 | 21 | 4.1% | −0.10 [−0.22, 0.02] |
| Chen X. et al. 2020 | 0.58 | 0.21 | 237 | 0.71 | 0.24 | 317 | 5.7% | −0.13 [−0.17, −0.09] |
| Chen X. et al. 2022 | 0.73 | 0.24 | 20 | 0.73 | 0.13 | 20 | 4.2% | 0.00 [−0.12, 0.12] |
| Das A. et al. 2020 | 0.94 | 0.2 | 22 | 0.81 | 0.18 | 28 | 4.4% | 0.13 [0.02, 0.24] |
| Liu et al. 2022 | 0.59 | 0.18 | 45 | 0.71 | 0.26 | 46 | 4.8% | −0.12 [−0.21, −0.03] |
| Liu Q. et al. 2021 | 0.9 | 0.16 | 42 | 0.99 | 0.06 | 42 | 5.5% | −0.09 [−0.14, −0.04] |
| Liu X. et al. 2022 | 0.6 | 0.2 | 33 | 0.71 | 0.3 | 21 | 3.6% | −0.11 [−0.26, 0.04] |
| Li W. et al. 2022 | 0.88 | 0.33 | 30 | 0.52 | 0.13 | 24 | 4.0% | 0.36 [0.23, 0.49] |
| Li X. et al. 2021 | 0.67 | 0.23 | 235 | 0.66 | 0.24 | 120 | 5.5% | 0.01 [−0.04, 0.06] |
| Miyajima K. et al. 2022 | 0.54 | 0.16 | 39 | 0.6 | 0.13 | 42 | 5.3% | −0.06 [−0.12, 0.00] |
| Niu H.X. et al. 2021 | 0.8 | 0.2 | 20 | 0.6 | 0.2 | 30 | 4.3% | 0.20 [0.09, 0.31] |
| Okubo Y. et al. 2022 | 0.6 | 0.2 | 43 | 0.8 | 0.3 | 46 | 4.5% | −0.20 [−0.31, −0.09] |
| Sharma P.S. et al. 2021 | 0.63 | 0.31 | 321 | 0.61 | 0.29 | 382 | 5.6% | 0.02 [−0.02, 0.06] |
| Sun Z. et al. 2020 | 0.78 | 0.19 | 16 | 0.71 | 0.19 | 16 | 3.9% | 0.07 [−0.06, 0.20] |
| Wang J.F. et al. 2020 | 0.94 | 0.21 | 61 | 0.92 | 0.15 | 65 | 5.3% | 0.02 [−0.04, 0.08] |
| Wang Z. et al. 2021 | 0.75 | 0.25 | 52 | 0.68 | 0.21 | 44 | 4.7% | 0.07 [−0.02, 0.16] |
| Zhang J.M. et al. 2019 | 0.68 | 0.2 | 20 | 0.51 | 0.06 | 21 | 4.8% | 0.17 [0.08, 0.26] |
| Zhang S. et al. 2021 | 0.68 | 0.22 | 19 | 0.73 | 0.23 | 37 | 4.1% | −0.05 [−0.17, 0.07] |
| Zhu H. et al. 2021 | 0.64 | 0.22 | 406 | 0.64 | 0.2 | 313 | 5.8% | 0.00 [−0.03, 0.03] |
| Total (95% CI) | | | 1764 | | | 1757 | 100.0% | 0.00 [−0.04, 0.05] |

Heterogeneity: $Tau^2 = 0.01$; $Chi^2 = 161.58$, df = 20 (P < 0.00001); $I^2 = 88\%$
Test for overall effect: Z = 0.18 (P = 0.86)

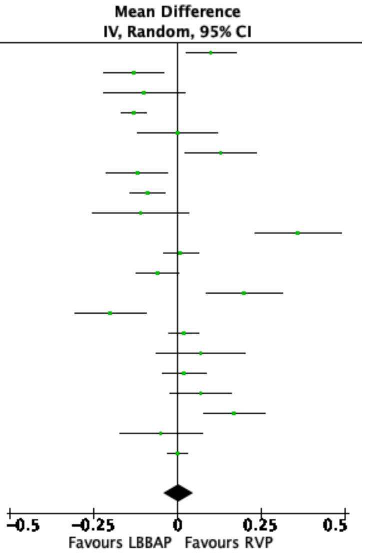

**Supplementary Figure S6:** Forest plot of ventricular impedance. (A) of LBBAP vs RVP group at implantation; (B) of LBBAP at implantation vs at follow-up; (C) of LBBAP vs RVP at follow-up.

A.

| Study or Subgroup | LBBAP Mean | LBBAP SD | LBBAP Total | RVP Mean | RVP SD | RVP Total | Weight | Mean Difference IV, Random, 95% CI |
|---|---|---|---|---|---|---|---|---|
| Byeon K. et al. 2022 | 645 | 103 | 42 | 729 | 165 | 84 | 6.5% | −84.00 [−131.07, −36.93] |
| Chen T.P. et al.2021 | 750.3 | 98.7 | 21 | 727.8 | 150.5 | 21 | 3.7% | 22.50 [−54.48, 99.48] |
| Chen X. et al. 2020 | 532.61 | 95.11 | 237 | 520.87 | 88.43 | 317 | 10.9% | 11.74 [−3.80, 27.28] |
| Chen X. et al. 2022 | 629.61 | 155.58 | 20 | 730.84 | 165.63 | 20 | 2.5% | −101.23 [−200.82, −1.64] |
| Das A. et al. 2020 | 703.27 | 31.54 | 22 | 681.82 | 42.2 | 28 | 10.3% | 21.45 [1.00, 41.90] |
| Liu Q. et al. 2021 | 752.38 | 108.44 | 42 | 730.95 | 69.49 | 42 | 7.6% | 21.43 [−17.52, 60.38] |
| Liu X. et al. 2022 | 830.61 | 120.6 | 33 | 817.14 | 104.98 | 21 | 5.0% | 13.47 [−47.43, 74.37] |
| Li W. et al. 2022 | 708.83 | 171.24 | 30 | 680 | 188.21 | 24 | 2.6% | 28.83 [−68.25, 125.91] |
| Li X. et al. 2021 | 757.2 | 164 | 235 | 853.6 | 258.5 | 120 | 6.0% | −96.40 [−147.18, −45.62] |
| Miyajima K. et al. 2022 | 644 | 114 | 39 | 614 | 128 | 42 | 5.8% | 30.00 [−22.71, 82.71] |
| Niu H.X. et al. 2021 | 696.2 | 124.7 | 20 | 686.3 | 110.8 | 30 | 4.4% | 9.90 [−57.62, 77.42] |
| Okubo Y. et al. 2022 | 575.5 | 85.9 | 43 | 610.7 | 120.8 | 46 | 7.0% | −35.20 [−78.53, 8.13] |
| Sun Z. et al. 2020 | 678.38 | 155.53 | 16 | 705.37 | 133.51 | 16 | 2.5% | −26.99 [−127.43, 73.45] |
| Wang J.F. et al. 2020 | 676.07 | 122.29 | 61 | 664.15 | 103.73 | 65 | 7.5% | 11.92 [−27.80, 51.64] |
| Wang Z. et al. 2021 | 791 | 146.2 | 52 | 793 | 161 | 44 | 4.9% | −2.00 [−63.98, 59.98] |
| Zhang J.M. et al. 2019 | 846.8 | 198.45 | 20 | 949.71 | 479.34 | 21 | 0.6% | −102.91 [−325.61, 119.79] |
| Zhang S. et al. 2021 | 818.83 | 165.73 | 29 | 844.29 | 182.95 | 37 | 3.2% | −25.46 [−109.80, 58.88] |
| Zhu H. et al. 2021 | 783 | 154 | 406 | 782 | 217 | 313 | 9.1% | 1.00 [−27.33, 29.33] |
| Total (95% CI) | | | 1368 | | | 1291 | 100.0% | −7.98 [−25.78, 9.83] |

Heterogeneity: $Tau^2 = 695.46$; $Chi^2 = 43.80$, df = 17 ($P = 0.0004$); $I^2 = 61\%$
Test for overall effect: Z = 0.88 (P = 0.38)

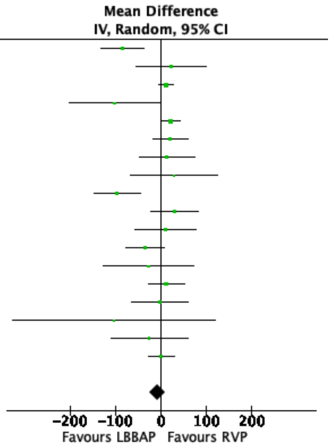


B.

| Study or Subgroup | LBBAP at Implantation Mean | LBBAP at Implantation SD | LBBAP at Implantation Total | LBBAP at FU Mean | LBBAP at FU SD | LBBAP at FU Total | Weight | Mean Difference IV, Random, 95% CI |
|---|---|---|---|---|---|---|---|---|
| Chen T.P. et al.2021 | 750.3 | 98.7 | 21 | 695.7 | 113.3 | 21 | 6.7% | 54.60 [−9.67, 118.87] |
| Chen X. et al. 2020 | 532.61 | 95.11 | 237 | 401.77 | 66.78 | 237 | 8.0% | 130.84 [116.04, 145.64] |
| Chen X. et al. 2022 | 629.61 | 155.58 | 20 | 455.78 | 73.68 | 20 | 6.3% | 173.83 [98.39, 249.27] |
| Das A. et al. 2020 | 703.27 | 31.54 | 22 | 714.55 | 137.09 | 22 | 6.9% | −11.28 [−70.06, 47.50] |
| Liu Q. et al. 2021 | 752.38 | 108.44 | 42 | 579.71 | 85.82 | 42 | 7.5% | 172.67 [130.85, 214.49] |
| Liu X. et al. 2022 | 830.61 | 120.6 | 33 | 583.59 | 69.87 | 33 | 7.3% | 247.02 [199.47, 294.57] |
| Li W. et al. 2022 | 708.83 | 171.24 | 30 | 624.93 | 80.65 | 27 | 6.6% | 83.90 [15.49, 152.31] |
| Li X. et al. 2021 | 757.2 | 164 | 235 | 621.3 | 149 | 235 | 7.8% | 135.90 [107.57, 164.23] |
| Miyajima K. et al. 2022 | 644 | 114 | 39 | 536 | 71 | 39 | 7.5% | 108.00 [65.85, 150.15] |
| Niu H.X. et al. 2021 | 696.2 | 124.7 | 20 | 507.6 | 72.3 | 20 | 6.8% | 188.60 [125.43, 251.77] |
| Okubo Y. et al. 2022 | 575.5 | 85.9 | 43 | 587.4 | 79.6 | 43 | 7.6% | −11.90 [−46.90, 23.10] |
| Wang J.F. et al. 2020 | 676.07 | 122.29 | 61 | 478.23 | 71.66 | 61 | 7.6% | 197.84 [162.27, 233.41] |
| Wang Z. et al. 2021 | 791 | 146.2 | 52 | 527.2 | 75.9 | 52 | 7.4% | 263.80 [219.03, 308.57] |
| Zhang S. et al. 2021 | 818.83 | 165.73 | 29 | 826.67 | 164.45 | 29 | 6.0% | −7.84 [−92.81, 77.13] |
| Total (95% CI) | | | 884 | | | 881 | 100.0% | 125.42 [85.01, 165.83] |

Heterogeneity: $Tau^2 = 5239.84$; $Chi^2 = 183.76$, df = 13 ($P < 0.00001$); $I^2 = 93\%$
Test for overall effect: Z = 6.08 (P < 0.00001)

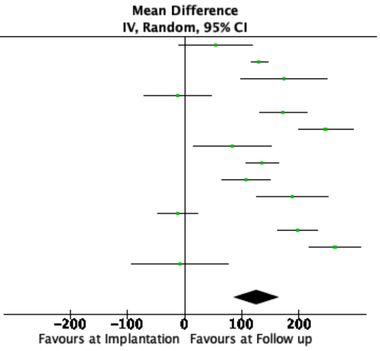


C.

| Study or Subgroup | LBBAP Mean | LBBAP SD | LBBAP Total | RVP Mean | RVP SD | RVP Total | Weight | Mean Difference IV, Random, 95% CI |
|---|---|---|---|---|---|---|---|---|
| Chen X. et al. 2020 | 401.77 | 66.78 | 237 | 492.11 | 58.97 | 317 | 9.4% | −90.34 [−101.04, −79.64] |
| Das A. et al. 2020 | 714.55 | 137.09 | 22 | 710.41 | 69.4 | 28 | 7.4% | 4.14 [−58.65, 66.93] |
| Liu Q. et al. 2021 | 579.71 | 85.82 | 42 | 537.29 | 90.59 | 42 | 8.6% | 42.42 [4.68, 80.16] |
| Liu X. et al. 2022 | 583.59 | 69.87 | 33 | 548.17 | 89.27 | 21 | 8.3% | 35.42 [−9.59, 80.43] |
| Li W. et al. 2022 | 624.93 | 80.65 | 27 | 606.17 | 90.31 | 23 | 8.1% | 18.76 [−29.07, 66.59] |
| Li X. et al. 2021 | 621.3 | 149 | 235 | 771.2 | 184.4 | 120 | 8.6% | −149.90 [−188.00, −111.80] |
| Miyajima K. et al. 2022 | 536 | 71 | 39 | 534 | 92 | 42 | 8.7% | 2.00 [−33.65, 37.65] |
| Niu H.X. et al. 2021 | 507.6 | 72.3 | 20 | 521 | 78.2 | 30 | 8.4% | −13.40 [−55.67, 28.87] |
| Okubo Y. et al. 2022 | 587.4 | 79.6 | 43 | 612.9 | 107.8 | 46 | 8.5% | −25.50 [−64.70, 13.70] |
| Wang J.F. et al. 2020 | 478.23 | 71.66 | 61 | 488.12 | 83.69 | 65 | 9.0% | −9.89 [−37.04, 17.26] |
| Wang Z. et al. 2021 | 527.2 | 75.9 | 52 | 529.6 | 98.5 | 44 | 8.7% | −2.40 [−38.07, 33.27] |
| Zhang S. et al. 2021 | 826.67 | 164.45 | 29 | 856.71 | 160.73 | 37 | 6.5% | −30.04 [−109.19, 49.11] |
| Total (95% CI) | | | 840 | | | 815 | 100.0% | −19.10 [−55.25, 17.05] |

Heterogeneity: $Tau^2 = 3594.47$; $Chi^2 = 154.36$, df = 11 ($P < 0.00001$); $I^2 = 93\%$
Test for overall effect: Z = 1.04 (P = 0.30)

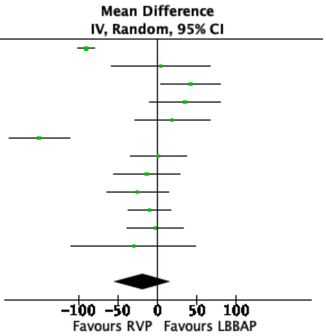

**Supplementary Figure S7:** Forest plot of R wave amplitude. (A) of LBBAP vs RVP group at implantation; (B) of LBBAP vs RVP at follow-up.

A.

| Study or Subgroup | LBBAP Mean | LBBAP SD | LBBAP Total | RVP Mean | RVP SD | RVP Total | Weight | Mean Difference IV, Random, 95% CI |
|---|---|---|---|---|---|---|---|---|
| Chen T.P. et al.2021 | 11.5 | 5.2 | 21 | 8.3 | 3.9 | 21 | 3.4% | 3.20 [0.42, 5.98] |
| Chen X. et al. 2020 | 11.82 | 4.24 | 237 | 11.21 | 3.32 | 317 | 9.1% | 0.61 [−0.04, 1.26] |
| Chen X. et al. 2022 | 9.43 | 4.14 | 20 | 9.32 | 2.19 | 20 | 4.9% | 0.11 [−1.94, 2.16] |
| Das A. et al. 2020 | 10.52 | 1.66 | 22 | 10.33 | 1.38 | 28 | 8.5% | 0.19 [−0.67, 1.05] |
| Liu X. et al. 2022 | 11.79 | 3.29 | 33 | 12.36 | 2.99 | 21 | 5.8% | −0.57 [−2.27, 1.13] |
| Li W. et al. 2022 | 10.93 | 5.74 | 30 | 13.68 | 6.62 | 24 | 2.6% | −2.75 [−6.10, 0.60] |
| Li X. et al. 2021 | 12.4 | 11.2 | 235 | 9.6 | 5.7 | 120 | 5.7% | 2.80 [1.04, 4.56] |
| Miyajima K. et al. 2022 | 12 | 5.7 | 39 | 11 | 4.5 | 42 | 4.4% | 1.00 [−1.25, 3.25] |
| Niu H.X. et al. 2021 | 11.2 | 3 | 20 | 11.5 | 3.4 | 30 | 5.6% | −0.30 [−2.09, 1.49] |
| Okubo Y. et al. 2022 | 17.4 | 5.6 | 43 | 11.6 | 3.9 | 46 | 5.0% | 5.80 [3.78, 7.82] |
| Sharma P.S. et al. 2021 | 11.34 | 5.63 | 321 | 10.54 | 5.08 | 382 | 8.7% | 0.80 [0.00, 1.60] |
| Sun Z. et al. 2020 | 12.37 | 3.65 | 16 | 9.81 | 4.08 | 16 | 3.6% | 2.56 [−0.12, 5.24] |
| Wang J.F. et al. 2020 | 12.16 | 3.64 | 61 | 11.46 | 3.46 | 65 | 7.2% | 0.70 [−0.54, 1.94] |
| Wang Z. et al. 2021 | 12.1 | 5.5 | 52 | 10.5 | 4.2 | 44 | 5.2% | 1.60 [−0.34, 3.54] |
| Zhang J.M. et al. 2019 | 9.28 | 5 | 20 | 11.03 | 3.14 | 21 | 3.8% | −1.75 [−4.32, 0.82] |
| Zhang S. et al. 2021 | 9.42 | 2.05 | 29 | 9.82 | 2.01 | 37 | 8.1% | −0.40 [−1.39, 0.59] |
| Zhu H. et al. 2021 | 11.7 | 6.1 | 406 | 10.6 | 4.9 | 313 | 8.6% | 1.10 [0.30, 1.90] |
| Total (95% CI) | | | 1605 | | | 1547 | 100.0% | 0.85 [0.21, 1.48] |

Heterogeneity: $Tau^2 = 1.04$; $Chi^2 = 54.27$, df = 16 ($P < 0.00001$); $I^2 = 71\%$
Test for overall effect: Z = 2.62 (P = 0.009)

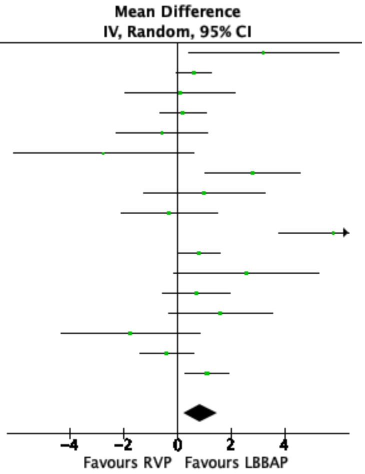


B.

| Study or Subgroup | LBBAP Mean | LBBAP SD | LBBAP Total | RVP Mean | RVP SD | RVP Total | Weight | Mean Difference IV, Random, 95% CI |
|---|---|---|---|---|---|---|---|---|
| Chen X. et al. 2020 | 13.86 | 5.99 | 237 | 11.58 | 3.44 | 317 | 8.9% | 2.28 [1.43, 3.13] |
| Das A. et al. 2020 | 10.32 | 1.39 | 22 | 10.12 | 1.61 | 28 | 9.0% | 0.20 [−0.63, 1.03] |
| Liu Q. et al. 2021 | 2.29 | 0.73 | 42 | 2.39 | 0.84 | 42 | 9.5% | −0.10 [−0.44, 0.24] |
| Liu X. et al. 2022 | 14.7 | 2.84 | 33 | 15.54 | 4.2 | 21 | 6.8% | −0.84 [−2.88, 1.20] |
| Li W. et al. 2022 | 16.88 | 7.47 | 30 | 17.86 | 7.56 | 24 | 3.7% | −0.98 [−5.02, 3.06] |
| Li X. et al. 2021 | 14.8 | 4.8 | 235 | 13 | 3.6 | 120 | 8.9% | 1.80 [0.91, 2.69] |
| Miyajima K. et al. 2022 | 15 | 4.9 | 39 | 12 | 4.6 | 42 | 6.7% | 3.00 [0.93, 5.07] |
| Niu H.X. et al. 2021 | 11 | 2.2 | 20 | 11.8 | 3.9 | 30 | 7.4% | −0.80 [−2.50, 0.90] |
| Okubo Y. et al. 2022 | 16.2 | 4.3 | 43 | 12.9 | 4.8 | 46 | 7.1% | 3.30 [1.41, 5.19] |
| Sharma P.S. et al. 2021 | 15.75 | 5.33 | 321 | 11.58 | 5.43 | 382 | 9.0% | 4.17 [3.37, 4.97] |
| Wang J.F. et al. 2020 | 14.78 | 4.7 | 61 | 14.15 | 3.59 | 65 | 7.9% | 0.63 [−0.84, 2.10] |
| Wang Z. et al. 2021 | 16.1 | 5.6 | 52 | 13.8 | 5.6 | 44 | 6.4% | 2.30 [0.05, 4.55] |
| Zhang S. et al. 2021 | 10.25 | 1.95 | 29 | 10.36 | 1.8 | 37 | 8.8% | −0.11 [−1.03, 0.81] |
| Total (95% CI) | | | 1164 | | | 1198 | 100.0% | 1.22 [0.23, 2.20] |

Heterogeneity: $Tau^2 = 2.62$; $Chi^2 = 137.51$, df = 12 ($P < 0.00001$); $I^2 = 91\%$
Test for overall effect: Z = 2.43 (P = 0.02)

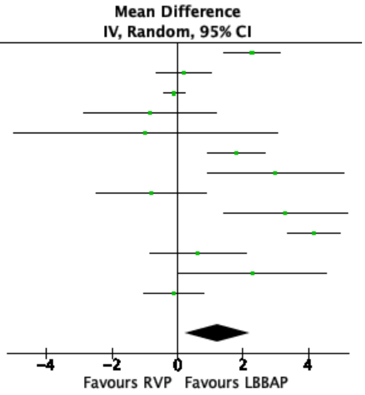

**Supplementary Figure S8.** Forest plots of. (A) procedural duration for LBBAP vs RVP group; (B) fluoroscopy time for LBBAP vs RVP group.

A.

| | LBBAP | | | RVP | | | | Mean Difference |
|---|---|---|---|---|---|---|---|---|
| Study or Subgroup | Mean | SD | Total | Mean | SD | Total | Weight | IV, Random, 95% CI |
| Das A. et al. 2020 | 63.15 | 7.02 | 22 | 55.15 | 6.16 | 28 | 11.9% | 8.00 [4.28, 11.72] |
| Liu et al. 2022 | 120.33 | 15.32 | 45 | 92.67 | 6.89 | 46 | 11.8% | 27.66 [22.76, 32.56] |
| Liu X. et al. 2022 | 120.82 | 11.46 | 33 | 93.19 | 8.7 | 21 | 11.7% | 27.63 [22.23, 33.03] |
| Li W. et al. 2022 | 158 | 48.98 | 30 | 152.08 | 55.64 | 24 | 6.6% | 5.92 [−22.41, 34.25] |
| Okubo Y. et al. 2022 | 130.2 | 22.6 | 43 | 110.4 | 14.3 | 46 | 11.3% | 19.80 [11.88, 27.72] |
| Sharma P.S. et al. 2021 | 97.7 | 32.65 | 321 | 71.8 | 31.82 | 382 | 11.8% | 25.90 [21.11, 30.69] |
| Wang J.F. et al. 2020 | 98.48 | 15.77 | 61 | 51.85 | 10.31 | 65 | 11.8% | 46.63 [41.95, 51.31] |
| Zhang J.M. et al. 2019 | 90.1 | 19.68 | 20 | 61.57 | 6.62 | 21 | 11.1% | 28.53 [19.45, 37.61] |
| Zhu H. et al. 2021 | 12.27 | 8.78 | 406 | 6.77 | 1.49 | 313 | 12.0% | 5.50 [4.63, 6.37] |
| Total (95% CI) | | | 981 | | | 946 | 100.0% | 22.44 [11.53, 33.36] |

Heterogeneity: $Tau^2 = 257.71$; $Chi^2 = 484.22$, df = 8 ($P < 0.00001$); $I^2 = 98\%$
Test for overall effect: Z = 4.03 ($P < 0.0001$)

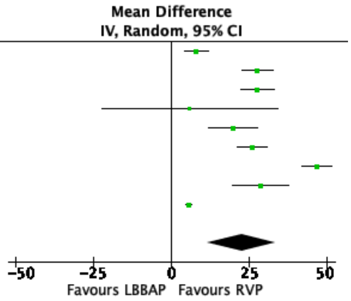


B.

| | LBBAP | | | RVP | | | | Mean Difference |
|---|---|---|---|---|---|---|---|---|
| Study or Subgroup | Mean | SD | Total | Mean | SD | Total | Weight | IV, Random, 95% CI |
| Chen X. et al. 2020 | 9.15 | 3.52 | 237 | 3.92 | 3.76 | 317 | 11.5% | 5.23 [4.62, 5.84] |
| Chen X. et al. 2022 | 7.92 | 4.92 | 20 | 0.83 | 0.8 | 20 | 10.5% | 7.09 [4.91, 9.27] |
| Das A. et al. 2020 | 6.08 | 1.42 | 22 | 5.06 | 1.3 | 28 | 11.5% | 1.02 [0.26, 1.78] |
| Li W. et al. 2022 | 9.13 | 2.89 | 30 | 7.25 | 2.42 | 24 | 11.1% | 1.88 [0.46, 3.30] |
| Okubo Y. et al. 2022 | 15.5 | 4.8 | 43 | 10.6 | 3.4 | 46 | 10.9% | 4.90 [3.16, 6.64] |
| Sharma P.S. et al. 2021 | 12.95 | 6.72 | 321 | 5.83 | 5.9 | 382 | 11.4% | 7.12 [6.18, 8.06] |
| Wang J.F. et al. 2020 | 16.07 | 3.61 | 61 | 3.58 | 1.32 | 65 | 11.4% | 12.49 [11.53, 13.45] |
| Zhang J.M. et al. 2019 | 15.55 | 5.62 | 20 | 4.67 | 2.06 | 21 | 10.1% | 10.88 [8.26, 13.50] |
| Zhu H. et al. 2021 | 5.3 | 3.72 | 406 | 2.73 | 1.19 | 313 | 11.6% | 2.57 [2.18, 2.96] |
| Total (95% CI) | | | 1160 | | | 1216 | 100.0% | 5.84 [3.49, 8.20] |

Heterogeneity: $Tau^2 = 12.47$; $Chi^2 = 511.44$, df = 8 ($P < 0.00001$); $I^2 = 98\%$
Test for overall effect: Z = 4.86 ($P < 0.00001$)

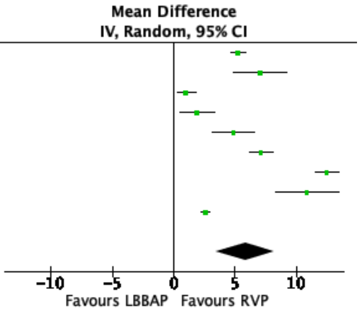

**Supplementary figure S9:** Funnel plots and Egger's test for all outcomes

A. QRSd native vs LBBAP, p = 0.004 (Egger).
B. QRSd LBBAP vs RVP at implantation p = 0.417 (Egger).
C. Intraventricular mechanical synchrony native vs LBBAP, p = 0.939 (Egger).
D. Intraventricular mechanical synchrony LBBAP vs RVP, p = 0.014 (Egger).
E. LVEF native vs LBBP, p = 0.918 (Egger).
F. LVEF LBBAP vs RVP, p = 0.573 (Egger).
G. LVEDD Native vs LBBAP, p = 0.339 (Egger).
H. LVEDD LBBAP vs RVP, p = 0.653 (Egger).
I. Pacing threshold LBBAP vs RVP post implantation, p = 0.218 (Egger).
J. Pacing threshold LBBAP post implantation vs at FU, p = 0.400 (Egger).
K. Ventricular impedance LBBAP vs RVP at implantation, p = 0.136 (Egger).
L. Ventricular impedance LBBAP at implantation vs at FU, p = 0.348 (Egger).
M. Ventricular impedance LBBAP vs RVP at FU, p = 0.318 (Egger).
N. R wave amplitude LBBAP vs RVP at implantation, p = 0.640 (Egger).
O. R wave amplitude LBBP vs RVP at FU, p = 0.728 (Egger).
P. Procedural duration LBBAP vs RVP, p = 0.701 (Egger).
Q. Fluoroscopy time LBBAP vs RVP, p = 0.194 (Egger).

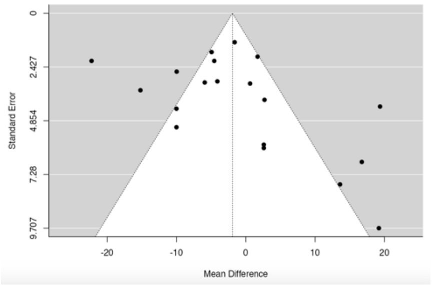


A

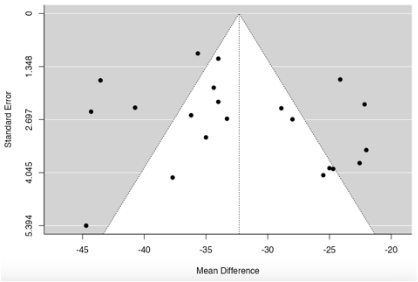


B

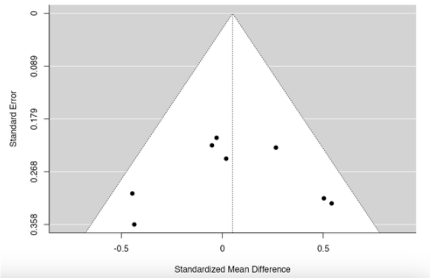


C

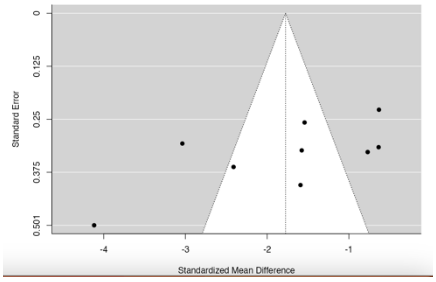


D

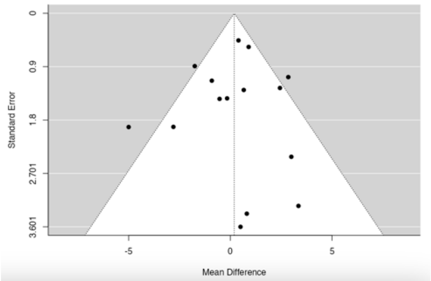
Standard Error
0
0.9
1.8
2.701
3.601
-5
0
5
Mean Difference

E

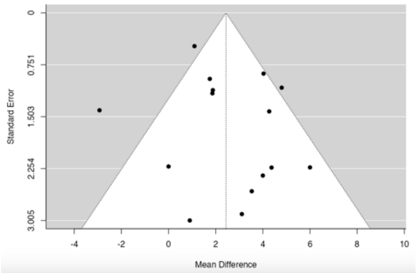
Standard Error
0
0.751
1.503
2.254
3.005
-4
-2
0
2
4
6
8
10
Mean Difference

F

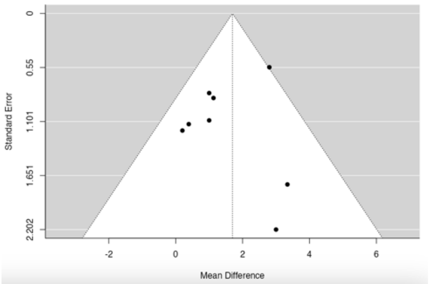
Standard Error
0
0.55
1.101
1.651
2.202
-2
0
2
4
6
Mean Difference

G

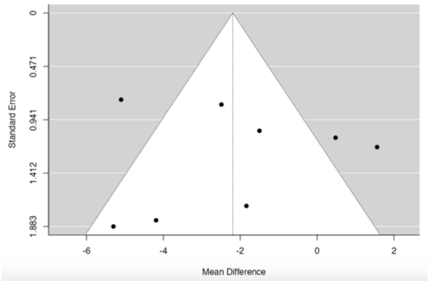
Standard Error
0
0.471
0.941
1.412
1.883
-6
-4
-2
0
2
Mean Difference

H

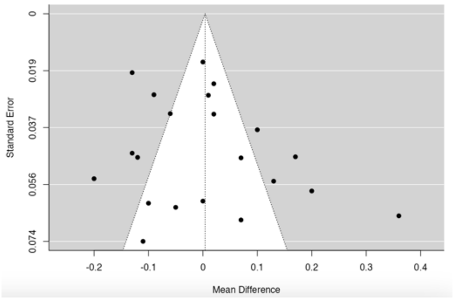
Standard Error
0
0.019
0.037
0.056
0.074
-0.2
-0.1
0
0.1
0.2
0.3
0.4
Mean Difference

I

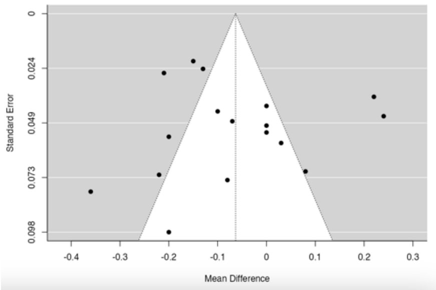
Standard Error
0
0.024
0.049
0.073
0.098
-0.4
-0.3
-0.2
-0.1
0
0.1
0.2
0.3
Mean Difference

J

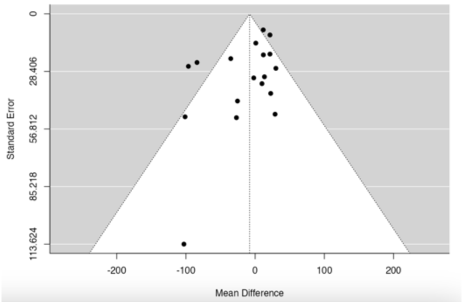
Standard Error
0
28.406
56.812
85.218
113.624
-200
-100
0
100
200
Mean Difference

K

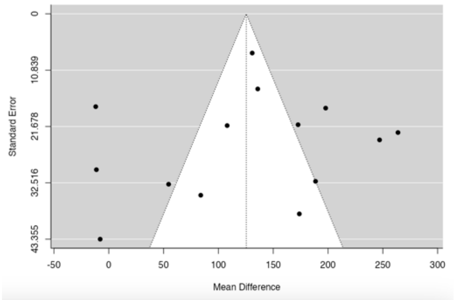
Standard Error
0
10.839
21.678
32.516
43.355
-50
0
50
100
150
200
250
300
Mean Difference

L

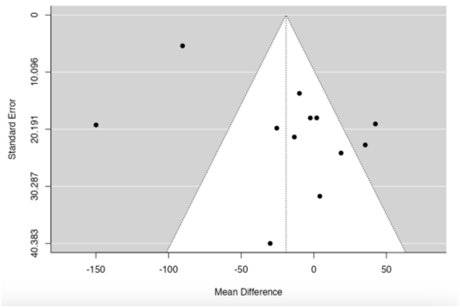
Standard Error
0
10.096
20.191
30.287
40.383
-150
-100
-50
0
50
Mean Difference

M

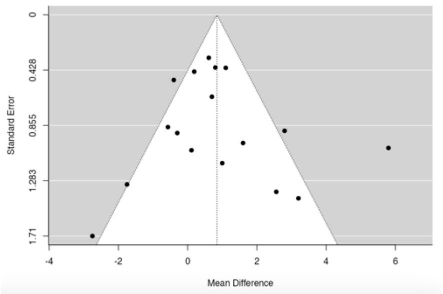
Standard Error
0
0.428
0.855
1.283
1.71
-4
-2
0
2
4
6
Mean Difference

N

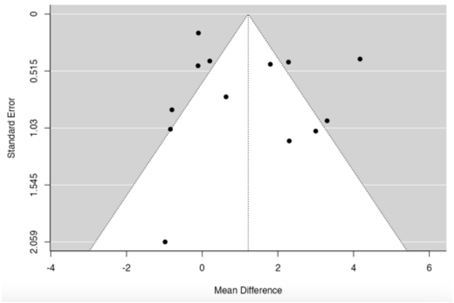
Standard Error
0
0.515
1.03
1.545
2.059
-4
-2
0
2
4
6
Mean Difference

O

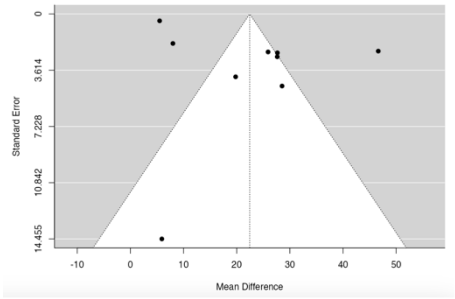
Standard Error
0
3.614
7.228
10.842
14.455
-10
0
10
20
30
40
50
Mean Difference

P

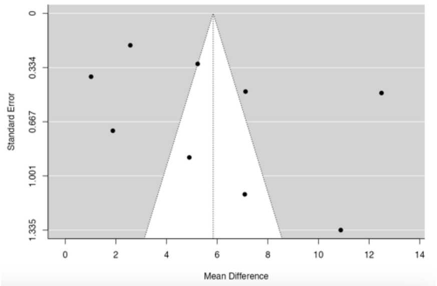
Standard Error
0
0.334
0.667
1.001
1.335
0
2
4
6
8
10
12
14
Mean Difference

Q
